# Clearance of asymptomatic genital HPV infections in young adult women is shaped by host immune response

**DOI:** 10.1101/2025.10.10.25337763

**Authors:** Inayat Bhardwaj, Vanina Boué, Tsukushi Kamiya, Christian Selinger, Massilva Rahmoun, Claire Bernat, Sophie Grasset, Soraya Groc, Marine Bonneau, Christelle Graf, Vincent Foulongne, Jacques Reynes, Vincent Tribout, Michel Segondy, Ignacio G Bravo, Nathalie Boulle, Carmen Lía Murall, Samuel Alizon, Nicolas Tessandier

## Abstract

Persistent infection with high-risk human papillomavirus (HPV) genotypes is an etiological factor in many cancers. The vast majority of HPV infections clear spontaneously within two years but the underlying mechanisms leading to this clearance remain unclear. Building on the PAPCLEAR longitudinal study, which followed young adult women with genital HPV infections, we analysed 100 vaginal swabs from 40 participants using quantitative bulk RNA sequencing to identify potential host transcriptomic signatures associated with infection clearance. Using a Gene Set Enrichment Analysis (GSEA) to detect significantly enriched pathways across infection outcome categories, we find that HPV-positive samples are characterised by a downregulation in both antiviral innate and adaptive immune responses. Independently of the genotype, non-clearing infections stand out by their adaptive immune response activation, which is surprisingly downregulated in persisting infections. This represents one of the rare transcriptomic analyses of asymptomatic HPV infection clearance. It stresses the key role of adaptive immunity, which represents a promising target for future diagnostics or immunotherapies.

**Author Summary:** *Study motivation:*

- Human papillomavirus (HPV) is one of the most prevalent sexually transmitted infections worldwide.
- Most HPV infections resolve spontaneously within about two years, but the mechanisms that drive natural clearance remain poorly understood.
- Persistent infection with certain high-risk HPV types can lead to precancerous lesions and, more rarely, cervical and other cancers.

*Study design and findings:*

- We used data from the PAPCLEAR study, a longitudinal cohort of young women with genital HPV infections, and analysed gene expression profiles from vaginal swabs collected from 40 participants.
- By comparing samples from cleared, persistent, and HPV-negative infections, we found that HPV positivity was associated with reduced expression of antiviral immune defences.
- Infections that did not clear showed stronger activation of adaptive immune pathways compared with clearing infections.

*Conclusions:*

- HPV-positive infections are associated with weakened antiviral immune responses.
- Non-clearing infections show activation of adaptive immunity.

## 1 Introduction

Human papillomaviruses (HPVs) are a diverse group of small DNA viruses that infect a range of epithelial sites in a tissue-specific manner [1]. Some genotypes, especially HPV16 and HPV18, are labelled as high-risk (HR) because of their well-established role in the development of cervical and other cancers [2, 3]. The vast majority of HPVs cause only benign or even asymptomatic infections in immunocompetent hosts [4, 5] and, according to longitudinal studies, more than 90 % of these infections are expected to be cleared within two years [6, 7]. However, some HPV infections persist and represent a major public health issue given their oncogenicity and high prevalence. For example, based on organised screening data, it is estimated that on average 3.2 to 4.7 % of the women aged 30 living in France at the end of 2023 were infected with HPV16 or HPV18 [8]. Despite the development of safe and efficient vaccines, HPV persistence is likely to remain a challenge for at least another generation because vaccine coverage is limited in many regions, especially in low and middle-income countries [9]. Furthermore, vaccine efficacy is lower when administered after a first infection [10, 11].

At the epidemiological level, multiple cofactors have been implicated in HPV infection persistence, which is the first step to precancerous lesions. These include high parity, a large number of sexual partners, host genetic susceptibility, smoking, or co-infection with other sexually transmitted pathogens such as herpes simplex virus type 2 (HSV-2) and *Chlamydia trachomatis* [12–17]. However, the causality in these associations is often unclear and our mechanistic understanding remains limited.

We know little about the biology of HPV infection clearance, or, more precisely, its becoming non-detectable [18]. Adaptive immunity could play a role, but we also know that at least half of the non-detection events do not lead to seroconversion and that natural immunity is poorly protective [19]. Innate immunity is also known to be involved in HPV cancers but there is little data for non-persisting infections in young adults [20]. Finally, stochasticity in the division of infected basal cells and the partitioning of viral episomes could also lead to infection clearance [21, 22].

Recently, the PAPCLEAR clinical study, which involved a high-resolution longitudinal follow-up of young women with genital HPV infections, provided new insights on the dynamics of HPV virus load over the course of an infection and its interaction with the immune response [23]. It showed that some immune populations, especially TCR-gamma-delta cells, are over-represented in HPV-positive cervical samples. Furthermore, it found the duration of the infection to be associated with specific patterns in terms of innate and adaptive immunity. However, this previous work mainly relied on local concentrations of cytokines [24] and on flow cytometry data from cervical smears, which limited its resolution.

Here, we further our understanding of host mechanisms associated with HPV clearance thanks to bulk RNA-Seq data from vaginal swabs collected during the same longitudinal study. We use log-linear models for differential gene expression and Gene Set Enrichment Analysis (GSEA) to identify significantly enriched pathways across infection outcomes, duration, multiplicity, and HPV genotypes. By comparing HPV-positive to HPV-negative samples, as well as samples from infections that become non-detectable to persisting ones, we identify host transcripts and pathways associated with HPV infections and their clearance.

## 2 Results

### 2.1 Cohort and dataset description

Among the 71 samples we could analyse after data pre-processing, the majority belonged to the ‘cleared’ (*n* = 27) or ‘not-cleared’ (*n* = 30) infection categories, while the rest (*n* = 14) originated from uninfected hosts (Fig 1a). The median follow-up duration was 193 days for the participants included in this analysis. Among the selected study samples and co-variates, ‘Vaccination Status’, ‘Intercourse with any sexual partner in the last two weeks’, ‘Menarche Age’, ‘BMI at the first visit’, and ‘Number of Lifetime Partners’ were the most significantly associated with the infection outcomes in a sample-level analysis (Table 1). These trends were expected based on the HPV literature (e.g., HPV16 samples all originated from non-vaccinated women or HPV-negative samples originated from women with fewer partners). Summary characteristics with a patient-level stratification can be found in Table S1.

**Table 1:** Characteristics of the *n* = 71 samples included in the analysis stratified by infection category. Values are shown as ‘(%)’ for categorical variables and ‘mean (Mean) and standard deviation (SD)’ and ‘Median and range (minimum-value, maximum-value)’ for continuous variables for samples in each category. Categorical comparisons were performed with Chi-squared test; continuous variables were compared with the Kruskal–Wallis rank-sum test to obtain unadjusted p-values.

| Characteristic | Cleared<br>(N=27) | Long, non-cleared<br>(N=7) | Negative<br>(N=14) | Not-cleared<br>(N=23) | P-value |
| --- | --- | --- | --- | --- | --- |
| **HPV vaccinated** |  |  |  |  |  |
| No | 10 (37.0%) | 3 (42.9%) | 8 (57.1%) | 23 (100%) | 0.001 |
| Yes | 17 (63.0%) | 4 (57.1%) | 6 (42.9%) | 0 (0%) |  |
| **Currently menstruating** |  |  |  |  |  |
| No | 20 (74.1%) | 5 (71.4%) | 9 (64.3%) | 15 (65.2%) | 0.924 |
| Yes | 7 (25.9%) | 2 (28.6%) | 4 (28.6%) | 8 (34.8%) |  |
| **Tobacco use** |  |  |  |  |  |
| No | 16 (59.3%) | 4 (57.1%) | 10 (71.4%) | 12 (52.2%) | 0.579 |
| Yes | 6 (22.2%) | 2 (28.6%) | 0 (0%) | 3 (13.0%) |  |
| Don't wish to answer | 4 (14.8%) | 1 (14.3%) | 4 (28.6%) | 3 (13.0%) |  |
| **Intercourse with regular partner** |  |  |  |  |  |
| No | 16 (59.3%) | 4 (57.1%) | 7 (50.0%) | 9 (39.1%) | 0.536 |
| Yes | 11 (40.7%) | 3 (42.9%) | 6 (42.9%) | 14 (60.9%) |  |
| **Self-reported stress** |  |  |  |  |  |
| 0 (Min) | 11 (40.7%) | 0 (0%) | 1 (7.1%) | 7 (30.4%) | 0.102 |
| 1 | 9 (33.3%) | 4 (57.1%) | 3 (21.4%) | 6 (26.1%) |  |
| 2 | 6 (22.2%) | 1 (14.3%) | 5 (35.7%) | 7 (30.4%) |  |
| 3 (Max) | 1 (3.7%) | 2 (28.6%) | 4 (28.6%) | 3 (13.0%) |  |
| **Intercourse with any partner, last 2 weeks** |  |  |  |  |  |
| No | 14 (51.9%) | 2 (28.6%) | 7 (50.0%) | 4 (17.4%) | 0.047 |
| Yes | 13 (48.1%) | 5 (71.4%) | 6 (42.9%) | 19 (82.6%) |  |
| **Age at first visit, years** |  |  |  |  |  |
| Mean (SD) | 20.5 (2.08) | 21.0 (2.16) | 20.1 (1.61) | 21.3 (1.71) | 0.226 |
| Median [Min, Max] | 19.0 [18.0, 24.0] | 21.0 [18.0, 24.0] | 19.0 [19.0, 23.0] | 21.0 [18.0, 25.0] |  |
| **Menarche age, years** |  |  |  |  |  |
| Mean (SD) | 12.4 (1.28) | 13.5 (1.22) | 12.5 (0.941) | 13.8 (1.17) | 0.001 |
| Median [Min, Max] | 12.0 [10.0, 14.0] | 14.0 [12.0, 15.0] | 12.5 [11.0, 14.0] | 14.0 [12.0, 16.0] |  |
| **Age at first intercourse, years** |  |  |  |  |  |
| Mean (SD) | 16.0 (1.63) | 17.0 (1.91) | 16.9 (1.17) | 16.7 (2.27) | 0.287 |
| Median [Min, Max] | 16.0 [13.0, 19.0] | 17.0 [15.0, 20.0] | 17.0 [15.0, 18.0] | 16.0 [14.0, 23.0] |  |
| **BMI at visit** |  |  |  |  |  |
| Mean (SD) | 23.1 (4.11) | 20.6 (1.46) | 22.1 (1.97) | 20.3 (2.56) | 0.03 |
| Median [Min, Max] | 23.2 [17.8, 31.4] | 21.1 [18.9, 23.1] | 21.7 [18.1, 25.2] | 19.5 [18.2, 25.9] |  |
| **Number of partners, lifetime mean** |  |  |  |  |  |
| Mean (SD) | 11.1 (12.3) | 14.4 (10.6) | 4.36 (1.98) | 21.1 (18.2) | 0.001 |
| Median [Min, Max] | 4.00 [2.00, 50.0] | 12.0 [3.00, 30.0] | 4.00 [1.00, 7.00] | 14.0 [3.00, 62.0] |  |

**Figure 1:**
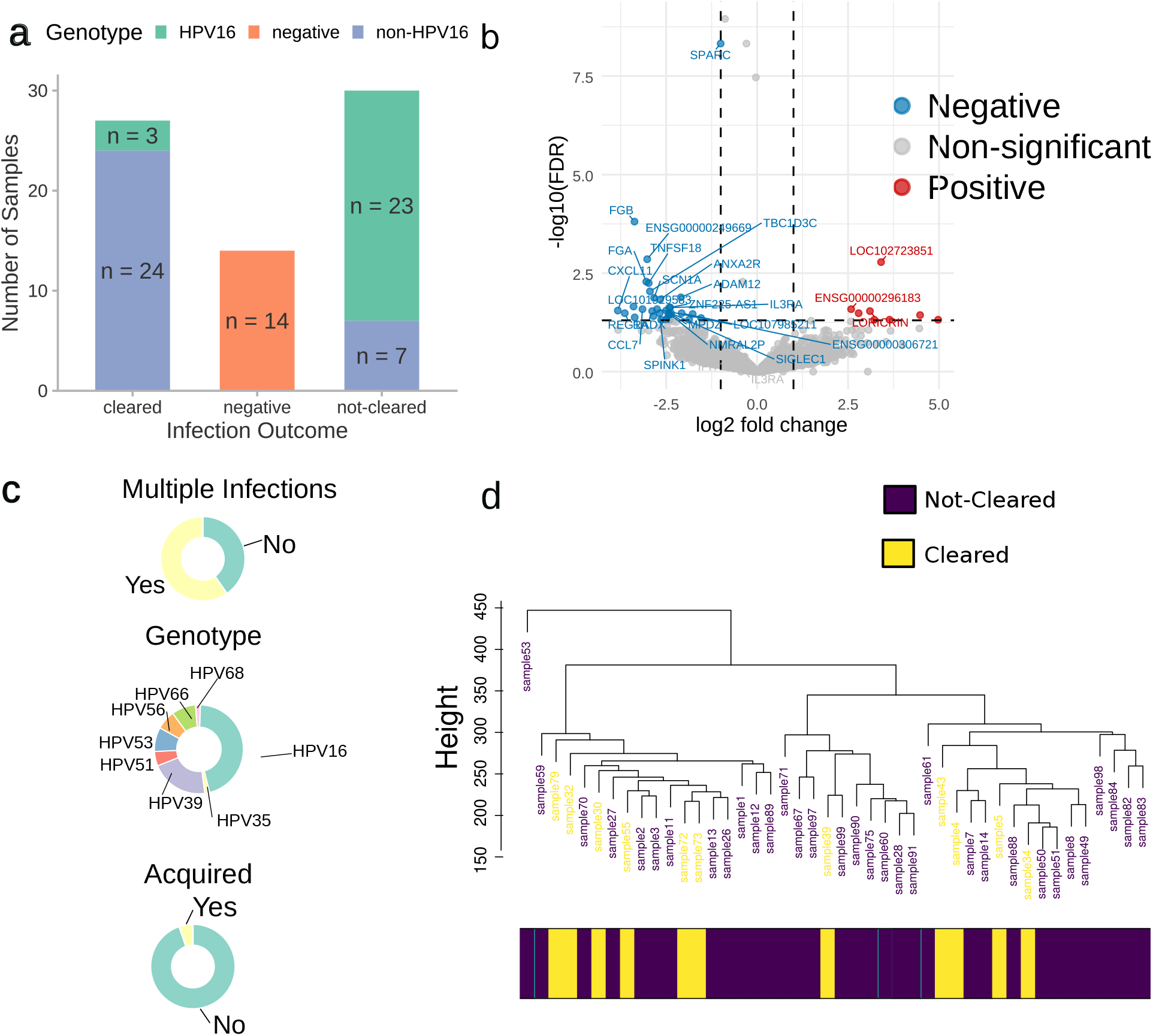
Sample characteristics and RNA global expression patterns in HPV infections. a) Samples origin stratified by infection outcome (clearance, no clearance, negative). b) Volcano-plot comparing gene expression in HPV-positive (*n* = 44) and HPV-negative samples (*n* = 27, the reference), with a log2-fold change cutoff of 1 and an adjusted p-value threshold of 0.05 (dashed lines). b) Distribution of HPV samples stratified by multiplicity of infection, HPV genotype, and whether the infection was acquired during the follow-up. d) Sample clustering dendrogram based on the gene co-expression matrices derived from the HPV-positive samples (*n* = 44) and stratified by infection status (‘cleared’ *vs*. ‘non-cleared’).

Importantly, a selection was made among the PAPCLEAR samples to focus on infection clearance or oncogenicity. Therefore, as shown in Fig 1b, non-clearing infections and the HPV16 genotype are over-represented compared to the whole cohort [7, 25]. Out of the 40 participants included in the study, only one had an acquired infection, *i*.*e*., her status changed from HPV-negative to HPV-positive during the follow-up. Finally, the majority of the infected participants (19 out of 32) were co-infected, *i*.*e*., infected by multiple HPV genotypes (Fig 1c).

A Differential Gene Expression (DEG) analysis across multiple comparisons in the full dataset only identified a handful of transcripts with an expression above the combined cut-offs in terms of fold change, FC, and false discovery rate, FDR (| log_2_(FC)| > 1 and FDR < 0.05). This is illustrated by Fig 1b when comparing HPV-positive (*n* = 44) to HPV-negative (*n* = 27) samples (the reference), with several genes, including CCL7, TNSF18, IL3RA, SIGLEC1, TBC1D3C and CXCL11 being significantly under-expressed. Focusing on HPV infections, there is an overlap in terms of gene expression between participants from the same category, *i*.*e*., clearing or non-clearing, as illustrated by the hierarchical clustering shown in Fig 1d.

### 2.2 HPV-positive samples exhibit immune downregulation

As a first analysis of the interaction between HPV infection and the host transcriptome, we compared HPV-positive (*n* = 44) to HPV-negative samples (*n* = 27) across all individuals in the study. As shown in Fig 2, we found several pathways to be downregulated in HPV-positive samples, including some associated with the adaptive immune response, *e*.*g*. pathways related to lymphocyte and leukocyte activation. Conversely, among the top 15 significantly regulated pathways, keratinisation and keratinocyte differentiation were the most positively enriched ones in HPV-positive samples.

**Figure 2:**
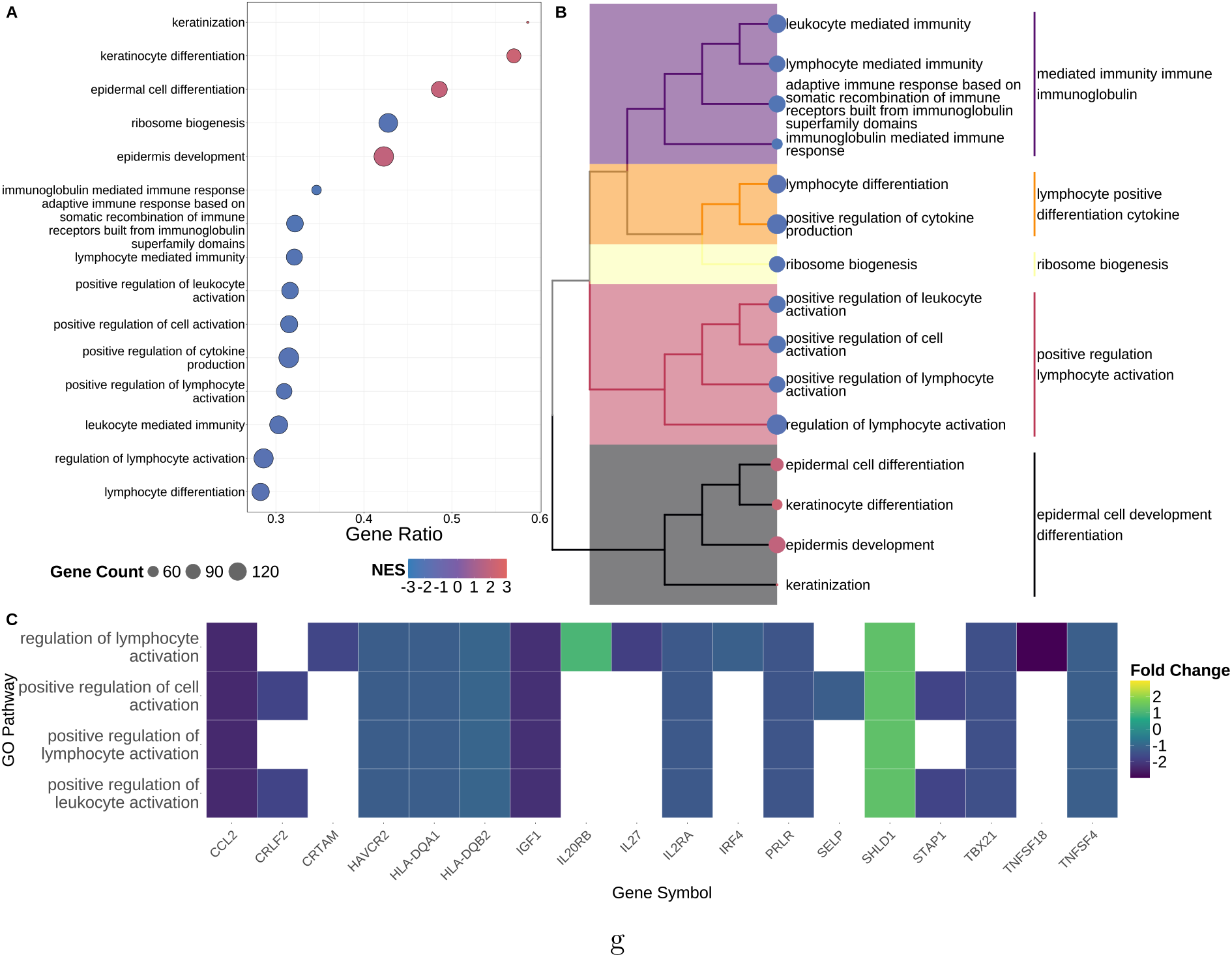
Gene Set Enrichment in HPV-positive *vs*. HPV-negative samples (the reference). a) Dot plot representing significantly positively-enriched (in red) or negatively-enriched (in blue) pathways across all HPV-positive (*n* = 44) *vs*. all HPV-negative samples (*n* = 27). Pathways are ranked by the ratio of genes involved (indicated by the gene ratio), with the dot size representing the gene set size in each pathway and the color the Normalised Enrichment Score (NES) of each pathway. b) Tree representing enriched gene sets clustered using Ward’s hierarchical clustering method based on the top 15 enriched gene sets obtained by comparing HPV-positive *vs*. HPV-negative samples. Each cluster represents a sub-tree containing the most related gene sets and is labelled using high-frequency terms related to the gene sets. c) Heatmap of the differential expression of genes overlapping in the pathway cluster linked to lymphocyte and leukocyte activation. Only overlapping genes with an absolute log fold change (FC) ≥ 2 are shown.

### 2.3 Innate immune response downregulation facilitates HPV-infections

Taking advantage of our sampling scheme and of the longitudinal nature of the dataset, we stratified samples based on infection status and outcome to compare all the samples from HPV infections (*n* = 57), to samples from individuals who remained uninfected throughout the study (*n* = 14). This comparative analysis identified pathways consistently down-regulated in HPV infections, especially some associated with antiviral immune responses—including defense response to virus, antiviral innate immune response, and response to type I interferon—and viral life cycle processes. Conversely, this analysis only found only two upregulated pathways, which corresponded to collagen fibril organisation and cell adhesion via plasma molecules, thereby highlighting the existence of potential virus-induced modulation of epithelial cell architecture (Fig 3). In terms of magnitude, the ratio of gene sets, i.e., the percentage of genes from a pathway that we find to be differentially expressed, varied from 0.2 to 0.5 in the significantly enriched pathways. At a finer scale, when focusing on differentially expressed genes, we found the most striking ones to be CCL8, IFIT1, CXCL11, and CXCL10, which were negatively down-regulated (p-value < 0.05) in HPV-infections compared to negatives (Fig S1a).

**Figure 3:**
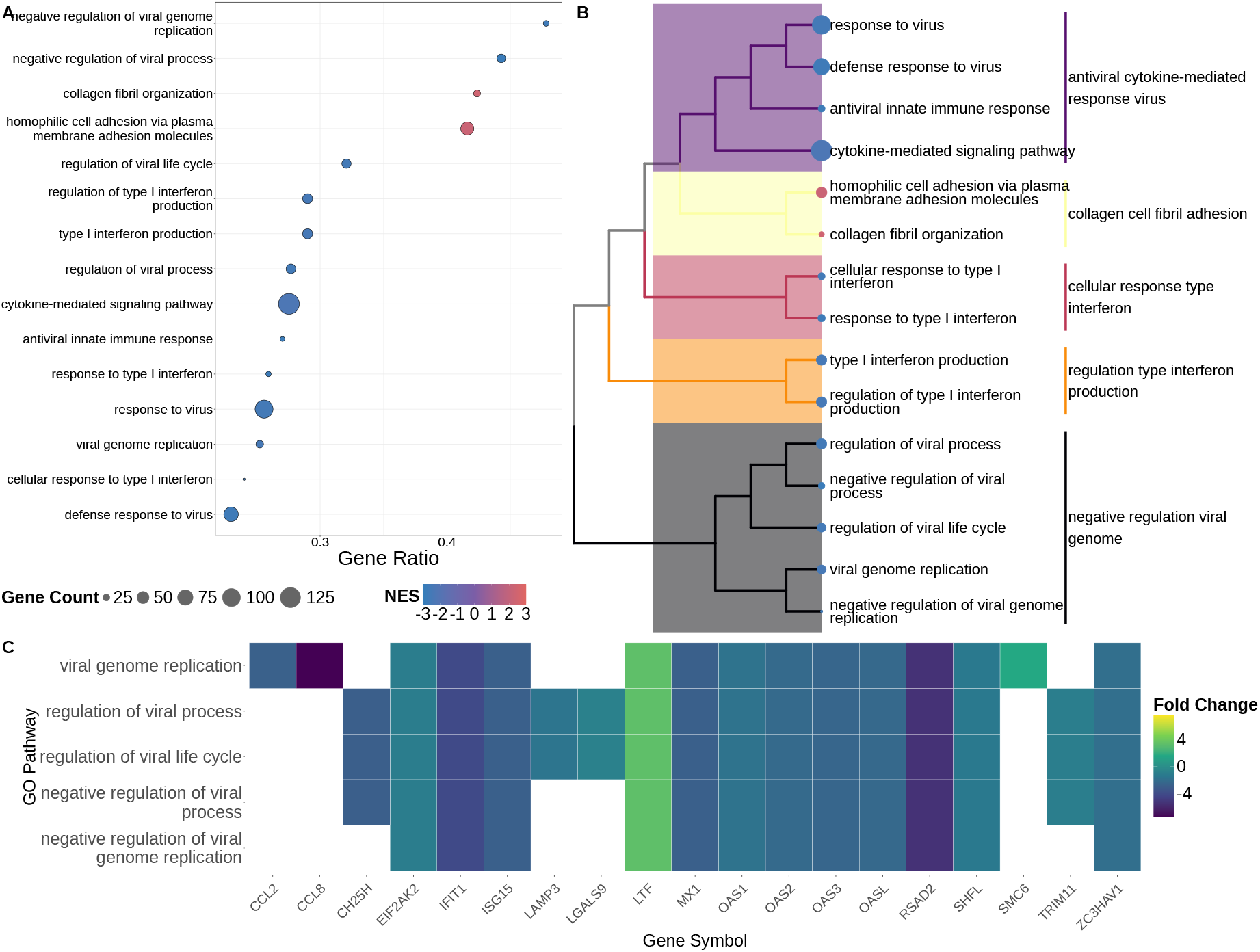
Gene Set Enrichment in samples from HPV infections *vs*. samples from uninfected hosts (the reference). a) Dotplot representing significantly positively and negatively enriched pathways in samples from HPV infections (*n* = 57) *vs*. samples from uninfected participants (*n* = 14). b) Tree representing enriched gene sets obtained by comparing HPV infections *vs*. uninfected participants. c) Heatmap of the differential expression of genes over-lapping in the pathway cluster linked to antiviral defense responses against viruses. See Fig 2 for details about the figure.

### 2.4 Cell differentiation and motility pathways characterise HPV16 infections

Approximately half of the HPV infection samples in our dataset originated from HPV16 infections (26 out of 57 samples). This sampling choice was made to investigate potential patterns associated with HPV16 infections, which is by far the most oncogenic one.

When comparing HPV16 (*n* = 26) and non-HPV16 (*n* = 31) infections, we did not find any differences in significantly enriched pathways related to immune responses (Fig. 4). However, pathways related to keratinisation, keratinocyte differentiation, and skin development were negatively enriched in HPV16 infections. The associated fold changes at the gene-expression level were low, suggesting a larger sample size may be needed to identify additional genotype-specific differences, as shown in Fig. S1b).

**Figure 4:**
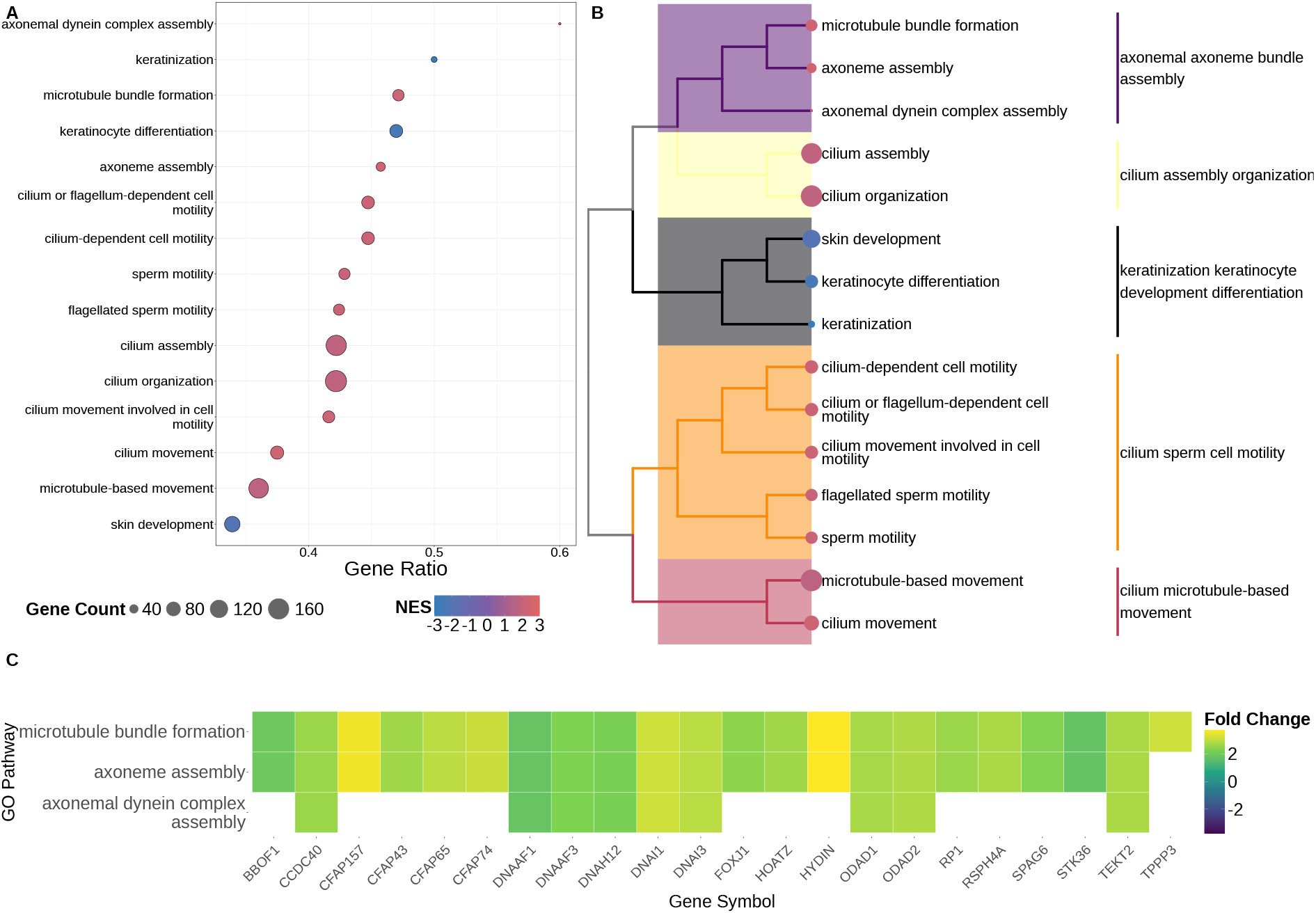
Gene Set Enrichment in samples from HPV16 infections *vs*. samples from non-HPV16 infections (the reference). a) Dotplot representing significantly positively and negatively enriched pathways in samples from HPV16 infections (*n* = 26) *vs*. samples from non-HPV16 infections (*n* = 31). b) Tree representing enriched gene sets obtained by comparing HPV infections *vs*. uninfected participants. c) Heatmap of the differential expression of genes overlapping in the pathway cluster linked to skin development, keratinocyte differentiation and cell motility. See Fig 2 for details about the figure.

### 2.5 Adaptive immune response is stronger in non-clearing infections

Moving to a stratification based on infection outcome, we compared the samples from infections that did not clear (*n* = 30 samples) to all the ones, i.e., HPV-positive and HPV-negative, originating from infections that did clear (*n* = 27 samples) by incroporating HPV-status (positive or negative) as a covariate in these analyses to account for the fact that infections, especially in the cleared category, which also included negative samples.

When analysing the clustering patterns of the enriched pathways in non-clearing infections, we found that adaptive-immunity modules dominated compared to the clearing ones (Fig 5b). In particular, immunoglobulin-mediated immune response, B-cell-mediated immunity, lymphocyte differentiation, and immune response activation and regulation, all displayed positive Normalised Enrichment Scores (NES ≈ +1.5 to +2.0), with gene ratios varying between 0.5 and 0.6. Furthermore, pathway enrichment in the non-clearing infections also revealed a strong negatively-enriched epithelial differentiation signature (Fig 5a). More precisely, ‘keratinization’, ‘epidermal/keratinocyte cell differentiation’, and ‘skin development’ pathways each exhibited gene ratios ranging from 0.3 to 0.4, with keratinization being the one with the highest proportion of gene sets involved amongst the negatively enriched pathways.

**Figure 5:**
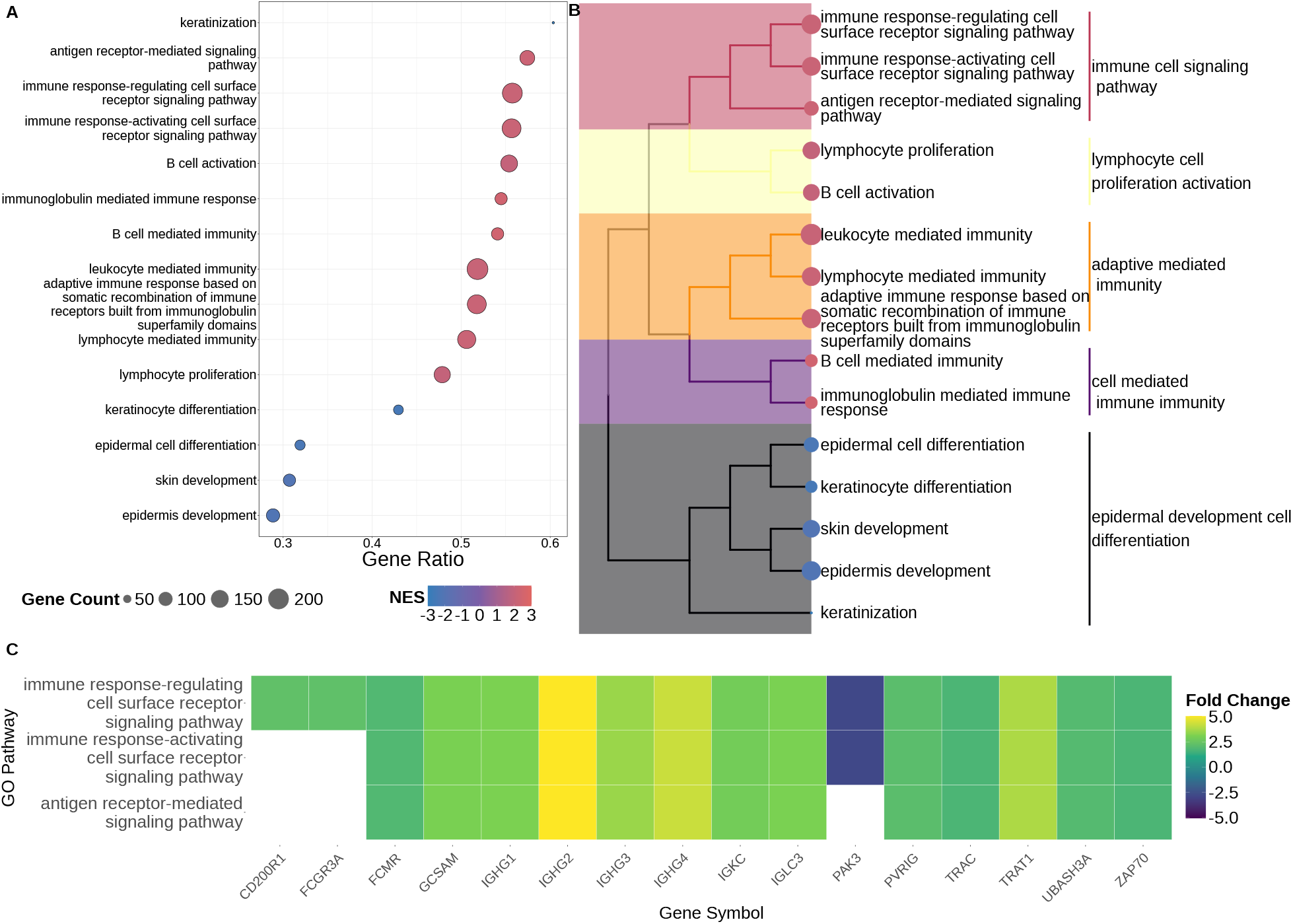
Gene Set Enrichment in samples from non-clearing *vs*. clearing infections (the reference). a) Dotplot representing significantly positively or negatively-enriched pathways across samples from non-clearing (*n* = 30) *vs*. clearing infections (*n* = 27). b) Tree representing enriched gene sets between non-clearing and clearing infection samples. c) Heatmap of the differential expression of genes overlapping in the pathway cluster linked to immune response activation and regulation. See Fig 2 for additional details about the figure.

**Figure 6:**
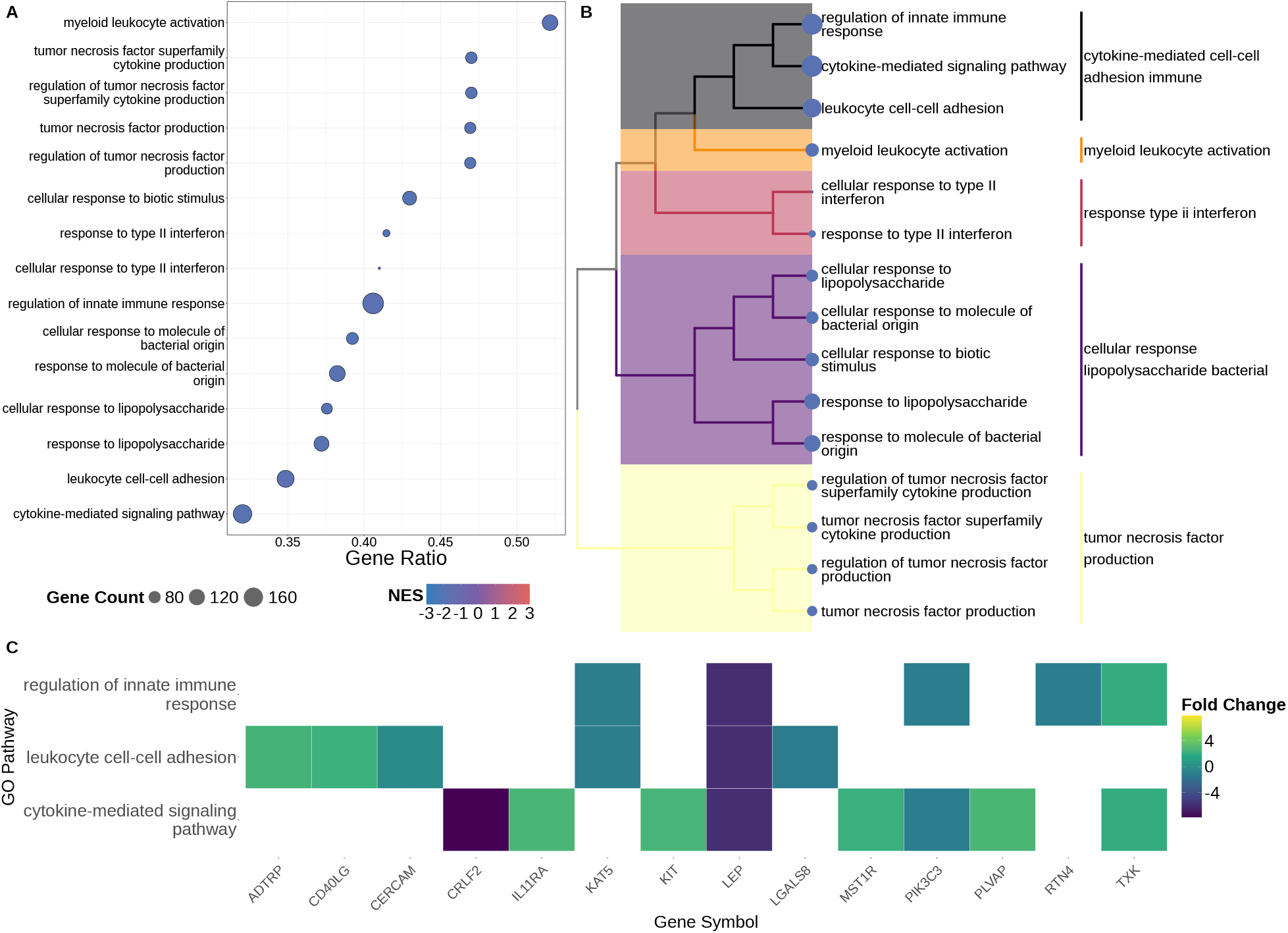
Gene Set Enrichment in samples from long non-clearing *vs*. shorter non-clearing infections (the reference). a) Dotplot representing significantly regulated pathways in samples from infections that were long, non-clearing (*n* = 7) *vs*. all samples from shorter non-clearing infections (*n* = 23). b) Tree representing enriched gene sets in the longer non-clearing vs short, non-clearing infection samples. c) Heatmap of the differential expression of genes overlapping in the pathway cluster linked to adaptive immunity mediated by lymphocytes, tumor necrosis factor, leukocytes, immunoglobulins and type B immune cells. See Figure 2 for details about the figure.

To check that our results were not confounded by the presence of multiple samples per participant, we analysed a subsampled dataset containing only the first sample (HPV-positive)from each infection (*i*.*e*., *n* = 16 non-clearing and *n* = 6 clearing samples). Given the decrease in statistical power, the analysis did not identify any significantly negatively enriched pathway. However, the most positively enriched pathways were related to immunoglobulin production and B-cell–mediated immune responses, followed by pathways involved in axoneme assembly, microtubule bundle formation, and cilium organisation (Supplementary Fig S4). Extracellular matrix and structural organisation pathways were also positively enriched.

At the level of the gene-transcript, in non-clearing infection samples, IGLV3-25 was the most significantly upregulated gene in non-clearing infections. This gene codes for an immunoglobulin that has been predicted to enable antigen-binding activity and is involved in immune responses (Fig. S1c).

### 2.6 Persistent infections are shaped by immune response modulation

Finally, we focused on samples from persistent infections (also referred to here as long non-clearing infections), which we assumed to be the ones lasting for at least 5 consecutive on-site visits, *i*.*e*. more than 10 months. When comparing samples from the long, non-cleared infections (*n* = 7) to the ones from short, non-clearing infections (*n* = 23), we found that they were associated with a downregulation of adaptive immune response pathways, with a normalised enrichment score (NES) of −1.78 to −2.0. Samples from persistent infections also showed a downregulation of B cell-mediated immunity, immunoglobulin-mediated immune responses, and pathways regulating leukocyte activation, which were already found on comparing all non-clearing infections to the clearing infections. Furthermore, the highest enrichment scores corresponded to interferon pathways and adaptive immune response pathways involving tumor necrosis and its regulation.

At the gene expression level, when comparing long non-clearing infections to shorter non-clearing infections, we found 49 DEG expressed genes with log_2_ FC > 1 and FDR < 0.05. The most significantly enriched genes belonged to immunoglobulin variants, including the IGLV2 and IGHV3 with up to five-fold increase in long-lasting samples (Fig. S1d).

## 3 Discussion

The vast majority of HPV infections become undetectable within two years. Independently of the debate as to whether or not this non-detection corresponds to actual clearance, *i*.*e*. whether the virus may persist with very low activity [18], its drivers remain poorly documented.

To address this question, we performed bulk transcriptomics analyses on vaginal samples that were collected longitudinally between 2016 and 2020 in Montpellier (France) in the PAP-CLEAR cohort [26]. Participants were young women aged between 18-25 years, with a majority of students [27]. During the follow-up, no high-grade lesions were detected and none of the participants vaccinated against high-risk HPVs developed an HPV16 infection, which is consistent with the demonstrated efficacy of these vaccines [25].

In the subset of the cohort we analysed, long-lasting HPV infections were associated with vaccination status, BMI, and average lifetime number of partners, which is consistent with the literature [6, 7, 18]. It has been previously shown that, besides age, most of the risk factors associated with contracting HPV infections are associated with sexual behaviour [28]. Note that in our dataset, none of the HPV16 infections persisted, although the concept of HPV persistence is currently being debated [18].

First, ignoring the longitudinal nature of our dataset, we compared HPV-positive to HPV-negative samples. This already allowed us to identify a downregulation of immune response pathways, some of which were associated with adaptive immune responses, e.g., activation and regulation of lymphocytes and leukocytes. This is consistent with the biology of HPV but is traditionally associated with chronic infections and cancers [1].

To further investigate the role of host-viral interactions in HPV clearance, we took advantage of the longitudinal PAPCLEAR follow-ups to classify HPV infections in terms of outcomes (‘clearing’ *vs*. ‘non-clearing’). For clearing infections, this meant by definition combining HPV-positive and HPV-negative samples in the same HPV infection class, but we accounted for HPV sample positivity in the analysis. Our control group included samples from participants who remained HPV-negative throughout their follow-up. We found that samples from HPV infections were associated with a strong downregulation of antiviral innate immune responses, including interferon signalling and defence responses to viral infection. Again, this echoes HPV behaviour that has been described in chronic infections or cancers and it is striking to detect it in acute infections in young adults.

One of our hypotheses was that HPV16 could be associated with different RNA expression patterns than other genotypes because of its striking difference in oncogenicity. A comparison between HPV16 infections and samples from uninfected hosts led to similar results as when including all the infections (Supplementary Fig S6). However, when comparing HPV16 infections to infections caused by genotypes, we did find differences with a decrease in several pathways, including keratinocyte differentiation in HPV16 samples. Again, this pattern is consistent with the known effect of the E6 protein of high-risk HPV genotypes [1] but it is striking to detect it in acute infections in young adults.

At finer resolution, when comparing samples from HPV infections to samples from uninfected participants, we detected several differentially expressed genes that had already been identified in previous studies. For example, CCL2, which is a key chemokine driving monocyte/macrophage recruitment processes, has been consistently associated with HPV infection progression to cancer [29, 30]. Another candidate for an HPV-specific marker that was differentially expressed in HPV infection samples, even HPV16 ones, is CXCL10. This cytokine, which can be used to identify potentially oncogenic HPV cases [31, 32], has been proposed as a potential biomarker for cervical cancer [33]. More recently, its concentration has been shown to be associated with shorter (asymptomatic) infections [23]. IFIT1 (interferon-induced protein with tetratricopeptide repeats 1) was also differentially expressed in HPV infections compared to uninfected participants and has been associated with replication inhibition in HPV infections [34]. In general, the pathways related to the host immune response against the virus were consistently downregulated in the HPV infection samples, whereas pathways related to the regulation of viral processes by the host innate immunity, including responses involving type I interferons, were consistently downregulated.

The density of the sampling process in the PAPCLEAR cohort allowed us to further stratify our samples based on infection outcome, namely ‘clearing’, ‘non-clearing’, or ‘long-non-clearing’. When comparing the latter, i.e., infections persisting for at least 10 months, to the ones that clear, we observed an upregulation of pathways associated with activation of the early adaptive immune response, which usually happens in the earlier stages of the infection [35]. This could result from either the innate immune response being unable to contain the infection or the adaptive immune response not being efficient in these infections, despite being activated. In the same comparative analysis, our gene set enrichment results also indicated a prominent induction of immunoglobulin genes (IGHV, IGHG, IGLV families). This immune profile is likely due to strong B cell maturation and activation and suggests that HPV persistence may be associated with a chronic, but potentially inefficient adaptive immune response.

In the shorter non-clearing infections, which were mostly caused by HPV16 in our dataset, we also observed an upregulation of immune responses associated with signalling, regulation, and mediation of adaptive immunity pathways, all of which are associated with the mounting of an adaptive immune response. However, there was also a significant downregulation of pathways associated with keratinisation, as well as keratinocyte and epidermal differentiation, which is consistent with the aforementioned specificity of HPV16 samples.

Furthermore, we found the adaptive immune response activation and regulation in persistent infections to be consistently downregulated as compared to shorter-non clearing infections. These findings, combined with our detection of a downregulation in tumor necrosis factor signalling, production, and regulation, indicate that even though adaptive immune responses are initiated during an HPV infection, these might be exhausted or inefficient at controlling the ongoing infection. However, these results need to be taken with caution since the majority of short non-clearing infections are caused by HPV16.

Most studies focus on chronic HPV infections or cancers [4] and most HPV cohorts typically have large intervals between observations [18]. This explains why, to the best of our knowledge, this is one of the first studies to investigate transcriptomic signatures of HPV clearance. More generally, few studies have attempted to characterise HPV-related host transcriptome changes. Some analyse tissues from HPV-induced cancers but, given that many cancer-associated variations in gene expression are likely to be independent of HPV-infection, these provide a limited opportunity to understand how HPV infections affect the host transcriptome. One exception is provided by a recent study that compared HPV-infected and uninfected samples from esophageal tissues [36]. They identified a differential expression of pathways linked to cell cycle and DNA replication in HPV-infected samples, which is comparable to the upregulation of pathways linked to keratinocyte and epidermal differentiation we identified in HPV-positive samples. Another relevant study used *in vitro* raft cultures to compare transcriptomic changes in high-risk HPV-infected epithelia [37]. This led them to identify an HPV-reprogrammed keratinocyte subpopulation, which further supports the role of HPV in modifying the keratinocyte environment it infects. Another study based on cell cultures performed bulk-transcriptomics and showed that infection with HPV16 was associated with alteration in the regulation of the viral life cycle, as well as keratinocyte differentiation pathways in immortalized human oral keratinocytes [38]. Again, these results are consistent with the changes we observe in HPV16-positive PAPCLEAR samples.

The vaginal microbiota composition is known to be associated with the risk of acquiring HPV infections [39, 40]. Therefore, we included it in all of our analyses as a covariate when estimating differential gene expression. To summarise its composition, we adopted the five community state types (CST) definition [41] because of its wide use in the field. This covariate was not significant in any of the analyses. However, when removing it from the model, we found that its presence was associated with the significance of related pathways, such as those linked to mucosal barrier integrity and immune-microbiota interactions (keratinocyte-related pathways). Furthermore, accounting for the CST further aided in distinguishing the features of non-clearing infections. This aligns with growing evidence that cervicovaginal microbiota composition could modulate HPV natural history [32, 42] and calls for more focused analyses of the transcriptomic differences across samples with varying vaginal microbiota composition.

Overall, our results provide an integrated view of the host response to HPV infection, linking transcriptomics changes to infection outcome. They identify candidate immune and epithelial processes that may underlie successful clearance or persistence. These results echo that from our recent study based on cytokine dosage and flow cytometry data from cervical smears, which concluded in hypothesising that either the innate immune response succeeds in rapidly clearing HPV infections, or an adaptive, but poorly efficient, immune response is mounted [23]. Thanks to the resolution of the transcriptomics data, we further identify the pathways involved in these host-virus interactions and even show that the adaptive immune response in the individuals who fail to clear the infection may be dysregulated (with a low expression of TNF). Future work integrating microbiome, proteomics, and immunological data will be critical to further dissect this interplay between the host, HPV, and the vaginal micro-environment in shaping infection trajectories.

## Material and methods

### Cohort design and stratification

The PAPCLEAR cohort included 189 women aged from 18 to 25 living in the area of Montpellier (France). After an inclusion visit and a results visit one month later, participants came back every 4 months if HPV-negative and every 2 months if HPV-positive [26]. They were followed until infection clearance or for a maximum of 24 months. HPV screening and genotyping was performed using the DEIA [43] and LIPA_25_ assays [44]. Clearance was defined as being negative for a specific genotype by LIPA_25_ detection for two consecutive visits, *i*.*e*., more than 2 months. Samples that remained positive for other alpha-papillomavirus detection via DEIA were considered to be ‘partially cleared’. We also define long non-clearing infections as infections with at least four consecutive detections of the same HPV genotype (Fig S2). Finally, participants who exhibited only transient HPV infections with ‘singletons’ (i.e., detection of a genotype at a single visit) were treated as ‘HPV-negative’.

Overall, we have three main categories related to HPV infections: cleared, non-cleared, and uninfected. Non-clearing infections can be further stratified into long-non-cleared and short-non-cleared (Supplementary Fig S2). Samples are stratified into three main classes: HPV16-positive, positive for another HPV genotype, and HPV-negative.

Bulk-RNA sequencing was used to analyse 100 PAPCLEAR samples that were selected to optimise four criteria: infection clearance, HPV16 positivity, long infections, and remaining uninfected. For all the associated visits, we also know the composition of the vaginal microbiota [45], which we here summarise using the vaginal microbiota type (CST) as a covariate.

### Sampling and sequencing

At the on-site clinical visits, a vaginal swab was performed by a gynaecologist or midwife using eSwabs^®^ (Copan Diagnostics, Murrieta, CA, USA) and was stored in 1 mL of an RNA preservation medium at −80°C.

Toal RNA extraction was done by the Genoscreen platform (Lille, France) with the NucleoSpin RNA Plus kit (Macherey-Nagel). RNA was quantified by RiboGreen fluorometry, and quality-checked on a Fragment Analyzer DNF-471 chip to validate integrity before library preparation. RNA was depleted using Illumina RiboPlus Depletion. Double-stranded cDNA was synthesized with random primers and RNase H, followed by end repair, Illumina adapter ligation, size selection with AMPure XP beads, and PCR amplification to generate sequencing libraries. The libraries were quantified by SYBR Green fluorimetry, size-checked by capillary electrophoresis (High Sensitivity kit; Fragment Analyzer DNF-474/Agilent HS), and further AMPure XP cleanups were performed to remove adapter dimers. Equimolar pooling was validated on a high-sensitivity DNA chip prior to sequencing.

Pooled libraries were sequenced on an Illumina platform in paired-end 2 *×* 150 bp mode. The run achieved Q30 ≥ 90.6% of bases (specification: ≥ 80%) and an average of ≈ 40.8 million paired reads per sample (the target being ≈ 30 million reads per sample).

### RNA-seq preprocessing and quantification

Raw RNA-seq reads were processed using the `nf-core/rnaseq` pipeline (version 3.14.0), a community-curated, scalable Nextflow workflow for transcriptome profiling [46]. The pipeline was executed with default settings unless specified and incorporated rigorous quality control, alignment, quantification, and reporting steps. Preprocessing involved the concatenation of FASTQ files, adapter trimming via `Trim Galore!`, and quality checks using `FastQC` [47]. Read strandedness was inferred using `Salmon`, and unique molecular identifiers (UMIs), if present, were extracted with `UMI-tools` [48]. Human reads were filtered using `BBsplit`.

Genome alignment and quantification was performed usiong the `STAR` aligner followed by transcript-level quantification using `Salmon` (quasi-mapping mode), following the default configuration of the pipeline [49, 50].

Post-alignment processing included deduplication (`UMI-tools`), sorting, indexing, and statistical summary generation using `SAMtools` [51], as well as duplicate marking (`Picard`). Transcript assembly was optionally performed with `StringTie` [52], and genome coverage tracks were generated using `BEDTools` [53] and `bedGraphToBigWig` [54].

Comprehensive quality control reports were generated using `MultiQC` [55], aggregating metrics from various stages. Additional QC modules included `dupRadar` [56], `Preseq` [57], `RSeQC` [58], and `Qualimap` [59] for assessing library complexity, duplication rates, and alignment quality.

### Gene expression filtering and normalization

Gene expression count matrixes were processed in R (v4.5) using the `edgeR` package (v3.42). Genes with low expression across all samples were removed using the `filterByExpr()` function. Only genes with a count higher than a minimum threshold of 10 counts per million and present in at least 70 % of the samples in the smallest group were considered. Library size normalization was performed using the trimmed mean of M-values (TMM) method. The resulting data were used to construct a specific model design matrix based on each stratification for downstream differential expression analysis.

### Differential gene expression analysis

A generalized linear model (GLM) with quasi-likelihood F-tests was applied to identify differentially expressed genes (DEGs) between `non-cleared` and `cleared` infection states, setting `cleared` as the reference level. Dispersion estimates were calculated using empirical Bayes shrinkage. Genes with a false discovery rate (FDR) below 0.05 were considered to be statistically significant.

### Gene Set Enrichment Analysis

Gene Set Enrichment Analysis (GSEA) was performed using the `clusterProfiler` package (v.4.10). Genes were ranked by multiplying the signed log_2_ fold change by the negative log_10_ of the p-value. The enrichment analysis was carried out using the `gseGO()` function for Gene Ontology (GO) Biological Process terms with parameters with an adjusted p-value cut off below 0.05 [60].

### Visualization and dimensionality reduction

The visualization of the Differential Gene Expression results was conducted with `ggplot2`. Volcano plots were generated using false discovery rate thresholds of 0.05 and | log_2_ FC| ≥ 1. Dot plots and cnetplots were used to represent enriched GO terms and their associated genes.

For global sample-level variation, t-distributed stochastic neighbour embedding (t-SNE) was applied to the log-transformed CPM values of the top 1,000 most variable genes using the `Rtsne` package with `dims = 3` and `perplexity = 10`. Samples were coloured with respect to their `RNAseq.group` classification to highlight transcriptomic substructure.

### Weighted Gene Co-expression Network Dendogram

Weighted Gene Co-expression Network Analysis (WGCNA) was performed using the `WGCNA` package in R to identify modules of co-expressed genes and explore transcriptomic patterns associated with infection outcomes. The log-transformed CPM matrix was transposed to generate a sample *×* gene matrix. Finally, hierarchical clustering was applied to detect and remove potential outlier samples.

## Data Availability

All data produced in the present study are available upon reasonable request to the authors

## Acknowledgements

The authors thank all the study participants, the CeGIDD and hospital staff from the CHU of Montpellier for their commitment and help. The authors acknowledge the ISO 9001-certified IRD i-Trop HPC facility, a member of the South Green Platform at IRD Montpellier, for providing high-performance computing resources used in this work. The authors thank Jacques Dainat at IRD Montpellier for his suggestions related to data pre-processing and analysis. This study was supported by the Expos’UM grant at Université de Montpellier to NT and the ANRS postdoctoral fellowship to IB.

## Supplementary Information

**Table S1:** Characteristics of the *N* = 40 participants included in the analysis stratified by infection category. See Table 1 for additional details.

|  | **cleared**<br>**(N=13)** | **HPV16**<br>**(N=12)** | **long.non.cleared**<br>**(N=7)** | **negative**<br>**(N=8)** | **p-**<br>**value** |
| --- | --- | --- | --- | --- | --- |
| HPV Vaccinated (%) |  |  |  |  |  |
| No | 4 (30.8%) | 12 (100%) | 3 (42.9%) | 5 (62.5%) | 0.004 |
| Yes | 9 (69.2%) | 0 (0%) | 4 (57.1%) | 3 (37.5%) |  |
| Currently Menstruating (%) |  |  |  |  |  |
| No | 8 (61.5%) | 5 (41.7%) | 5 (71.4%) | 5 (62.5%) | 0.582 |
| Yes | 5 (38.5%) | 7 (58.3%) | 2 (28.6%) | 3 (37.5%) |  |
| Tobacco Use (%) |  |  |  |  |  |
| Don't wish to answer | 2 (15.4%) | 3 (25.0%) | 1 (14.3%) | 3 (37.5%) | 0.745 |
| No | 8 (61.5%) | 7 (58.3%) | 4 (57.1%) | 5 (62.5%) |  |
| Yes | 3 (23.1%) | 2 (16.7%) | 2 (28.6%) | 0 (0%) |  |
| Intercourse with regular partner (%) |  |  |  |  |  |
| No | 6 (46.2%) | 2 (16.7%) | 4 (57.1%) | 4 (50.0%) | 0.249 |
| Yes | 7 (53.8%) | 10 (83.3%) | 3 (42.9%) | 4 (50.0%) |  |
| Self-Reported Stress (%) |  |  |  |  |  |
| 3 (Max) | 0 (0%) | 1 (8.3%) | 2 (28.6%) | 2 (25.0%) | 0.235 |
| 1 | 3 (23.1%) | 4 (33.3%) | 4 (57.1%) | 3 (37.5%) |  |
| 0 (Min) | 7 (53.8%) | 4 (33.3%) | 0 (0%) | 1 (12.5%) |  |
| 2 | 3 (23.1%) | 3 (25.0%) | 1 (14.3%) | 1 (12.5%) |  |
| Intercourse with any partner (last 2 weeks) (%) |  |  |  |  |  |
| No | 4 (30.8%) | 0 (0%) | 2 (28.6%) | 4 (50.0%) | 0.074 |
| Yes | 9 (69.2%) | 12 (100%) | 5 (71.4%) | 4 (50.0%) |  |
| Age at First Visit (years) |  |  |  |  |  |
| Mean (SD) | 20.8 (2.42) | 21.3 (2.02) | 21.0 (2.16) | 20.0 (1.60) | 0.577 |
| Median [Min, Max] | 19.0 [18.0, 24.0] | 21.0 [18.0, 25.0] | 21.0 [18.0, 24.0] | 19.0 [19.0, 23.0] |  |
| Menarche Age (years) |  |  |  |  |  |
| Mean (SD) | 12.2 (1.48) | 13.8 (1.27) | 13.5 (1.22) | 12.5 (0.926) | 0.031 |
| Median [Min, Max] | 12.0 [10.0, 14.0] | 14.0 [12.0, 16.0] | 14.0 [12.0, 15.0] | 12.5 [11.0, 14.0] |  |
| Age at First Intercourse (years) |  |  |  |  |  |
| Mean (SD) | 15.9 (1.71) | 16.6 (2.31) | 17.0 (1.91) | 17.0 (1.20) | 0.393 |
| Median [Min, Max] | 16.0 [13.0, 19.0] | 16.0 [14.0, 23.0] | 17.0 [15.0, 20.0] | 17.5 [15.0, 18.0] |  |
| BMI at Visit |  |  |  |  |  |
| Mean (SD) | 23.7 (4.69) | 21.0 (2.66) | 20.6 (1.46) | 21.7 (2.19) | 0.434 |
| Median [Min, Max] | 23.2 [17.8, 31.4] | 20.7 [18.2, 25.9] | 21.1 [18.9, 23.1] | 21.5 [18.1, 25.2] |  |
| Number of Partners (lifetime mean) |  |  |  |  |  |
| Mean (SD) | 12.6 (14.6) | 19.4 (17.2) | 14.4 (10.6) | 4.13 (2.10) | 0.02 |
| Median [Min, Max] | 4.00 [2.00, 50.0] | 14.0 [3.00, 62.0] | 12.0 [3.00, 30.0] | 4.00 [1.00, 7.00] |  |

**Table S2:**
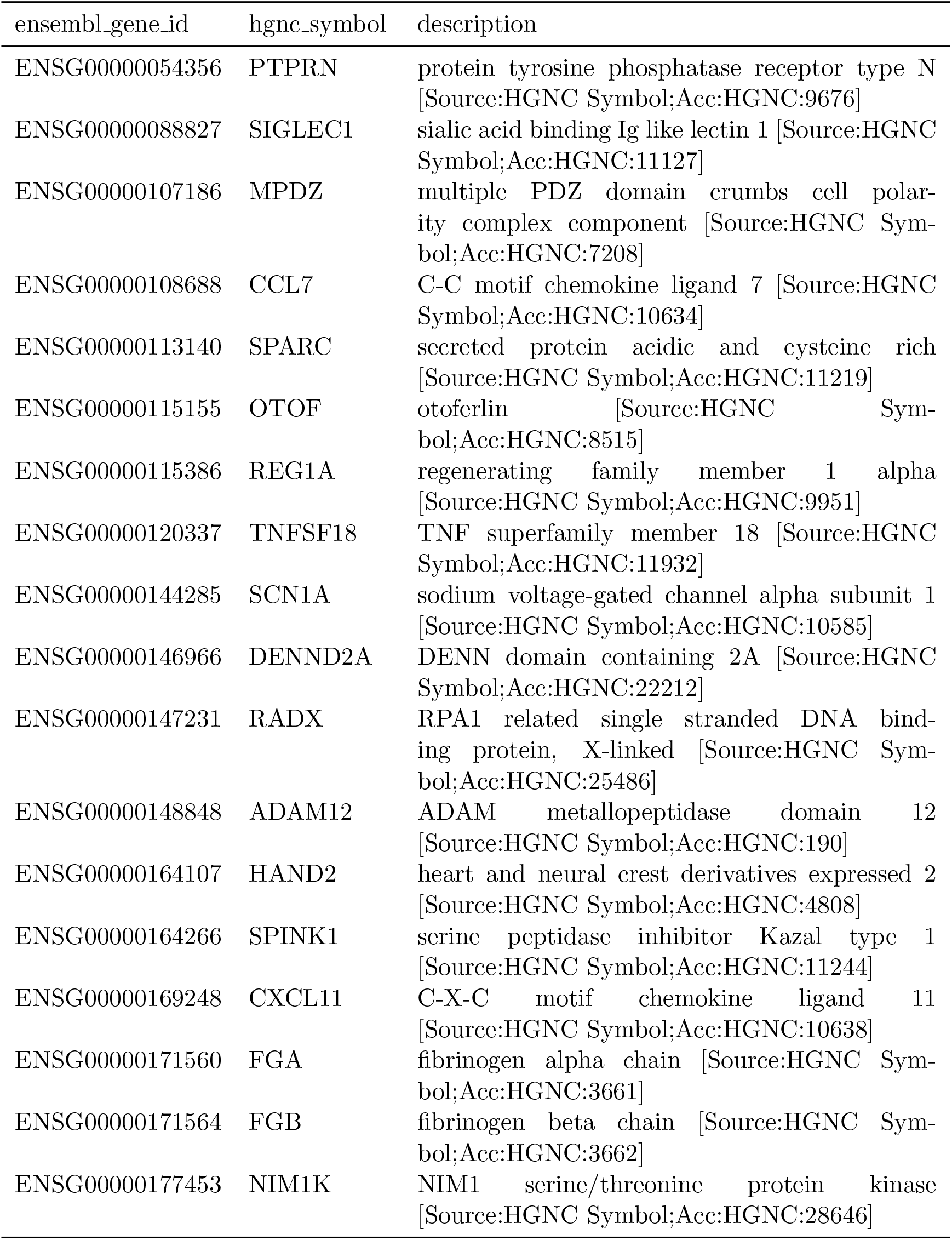
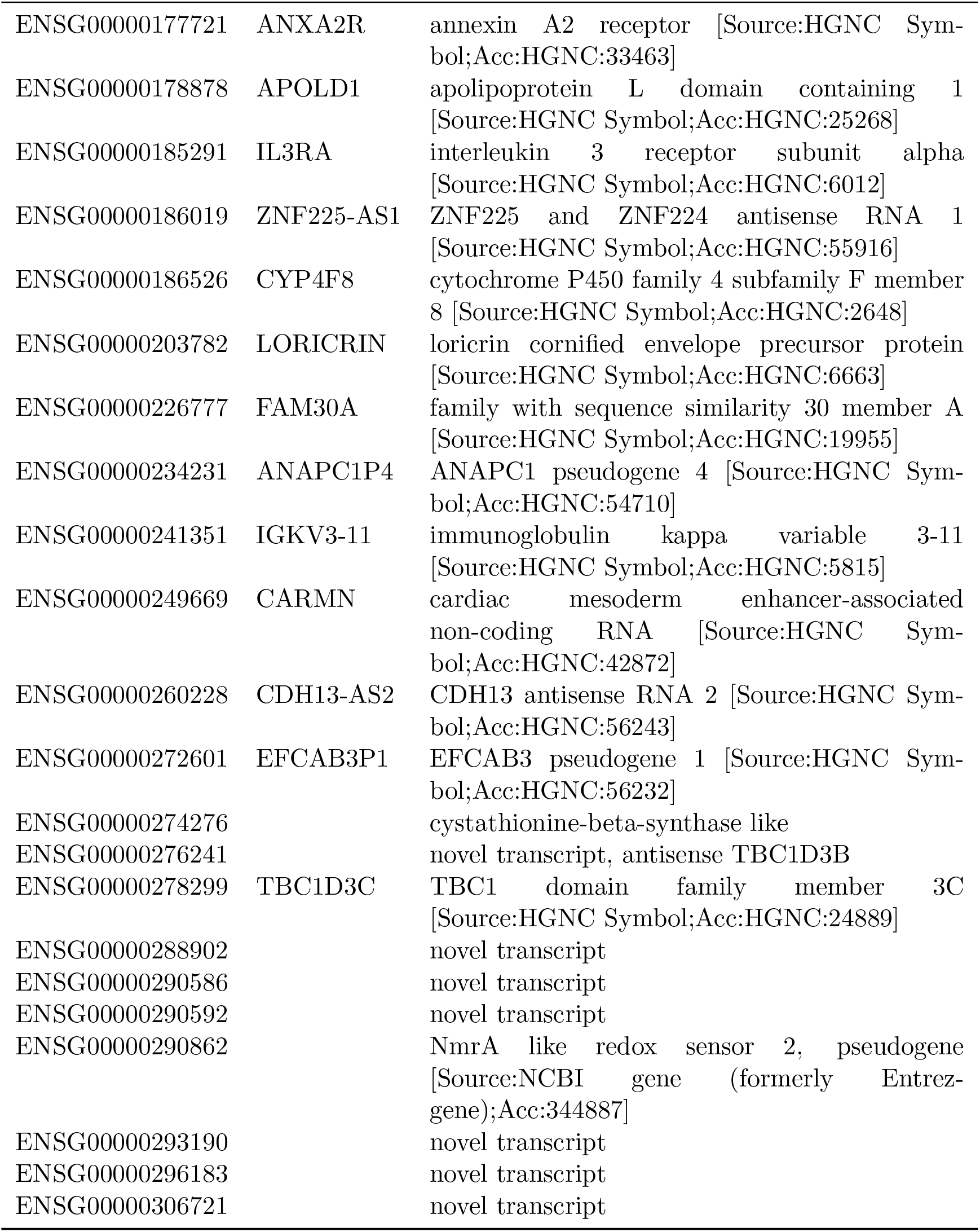
Annotation of the 40 input Ensembl genes across HPV-positive and HPV-negative samples.

**Table S3:** Annotation of the 69 input Ensembl genes differentially expressed between HPV vs uninfected hosts.

| ensembl_gene_id | hgnc_symbol | description |
| --- | --- | --- |
| ENSG00000002549 | LAP3 | leucine aminopeptidase 3 [Source:HGNC Symbol;Acc:HGNC:18449] |
| ENSG00000017427 | IGF1 | insulin like growth factor 1 [Source:HGNC Symbol;Acc:HGNC:5464] |
| ENSG00000077274 | CAPN6 | calpain 6 [Source:HGNC Symbol;Acc:HGNC:1483] |
| ENSG00000079931 | MOXD1 | monooxygenase DBH like 1 [Source:HGNC Symbol;Acc:HGNC:21063] |
| ENSG00000102794 | ACOD1 | aconitate decarboxylase 1 [Source:HGNC Symbol;Acc:HGNC:33904] |
| ENSG00000104332 | SFRP1 | secreted frizzled related protein 1 [Source:HGNC Symbol;Acc:HGNC:10776] |
| ENSG00000105939 | ZC3HAV1 | zinc finger CCCH-type containing, antiviral 1 [Source:HGNC Symbol;Acc:HGNC:23721] |
| ENSG00000106483 | SFRP4 | secreted frizzled related protein 4 [Source:HGNC Symbol;Acc:HGNC:10778] |
| ENSG00000108700 | CCL8 | C-C motif chemokine ligand 8 [Source:HGNC Symbol;Acc:HGNC:10635] |
| ENSG00000108821 | COL1A1 | collagen type I alpha 1 chain [Source:HGNC Symbol;Acc:HGNC:2197] |
| ENSG00000111331 | OAS3 | 2'-5'-oligoadenylate synthetase 3 [Source:HGNC Symbol;Acc:HGNC:8088] |
| ENSG00000111335 | OAS2 | 2'-5'-oligoadenylate synthetase 2 [Source:HGNC Symbol;Acc:HGNC:8087] |
| ENSG00000113361 | CDH6 | cadherin 6 [Source:HGNC Symbol;Acc:HGNC:1765] |
| ENSG00000115155 | OTOF | otoferlin [Source:HGNC Symbol;Acc:HGNC:8515] |
| ENSG00000119917 | IFIT3 | interferon induced protein with tetratricopeptide repeats 3 [Source:HGNC Symbol;Acc:HGNC:5411] |
| ENSG00000119922 | IFIT2 | interferon induced protein with tetratricopeptide repeats 2 [Source:HGNC Symbol;Acc:HGNC:5409] |
| ENSG00000120645 | IQSEC3 | IQ motif and Sec7 domain ArfGEF 3 [Source:HGNC Symbol;Acc:HGNC:29193] |
| ENSG00000122786 | CALD1 | caldesmon 1 [Source:HGNC Symbol;Acc:HGNC:1441] |

*Table S3 continued from previous page*
| ensembl_gene_id | hgnc_symbol | description |
| --- | --- | --- |
| ENSG00000123243 | ITIH5 | inter-alpha-trypsin inhibitor heavy chain 5 [Source:HGNC Symbol;Acc:HGNC:21449] |
| ENSG00000129009 | ISLR | immunoglobulin superfamily containing leucine rich repeat [Source:HGNC Symbol;Acc:HGNC:6133] |
| ENSG00000134321 | RSAD2 | radical S-adenosyl methionine domain containing 2 [Source:HGNC Symbol;Acc:HGNC:30908] |
| ENSG00000134326 | CMPK2 | cytidine/uridine monophosphate kinase 2 [Source:HGNC Symbol;Acc:HGNC:27015] |
| ENSG00000135114 | OASL | 2'-5'-oligoadenylate synthetase like [Source:HGNC Symbol;Acc:HGNC:8090] |
| ENSG00000143341 | HMCN1 | hemicentin 1 [Source:HGNC Symbol;Acc:HGNC:19194] |
| ENSG00000144057 | ST6GAL2 | ST6 beta-galactoside alpha-2,6-sialyltransferase 2 [Source:HGNC Symbol;Acc:HGNC:10861] |
| ENSG00000144285 | SCN1A | sodium voltage-gated channel alpha subunit 1 [Source:HGNC Symbol;Acc:HGNC:10585] |
| ENSG00000150471 | ADGRL3 | adhesion G protein-coupled receptor L3 [Source:HGNC Symbol;Acc:HGNC:20974] |
| ENSG00000152402 | GUCY1A2 | guanylate cyclase 1 soluble subunit alpha 2 [Source:HGNC Symbol;Acc:HGNC:4684] |
| ENSG00000154096 | THY1 | Thy-1 cell surface antigen [Source:HGNC Symbol;Acc:HGNC:11801] |
| ENSG00000157601 | MX1 | MX dynamin like GTPase 1 [Source:HGNC Symbol;Acc:HGNC:7532] |
| ENSG00000158270 | COLEC12 | collectin subfamily member 12 [Source:HGNC Symbol;Acc:HGNC:16016] |
| ENSG00000164107 | HAND2 | heart and neural crest derivatives expressed 2 [Source:HGNC Symbol;Acc:HGNC:4808] |
| ENSG00000164742 | ADCY1 | adenylate cyclase 1 [Source:HGNC Symbol;Acc:HGNC:232] |
| ENSG00000169245 | CXCL10 | C-X-C motif chemokine ligand 10 [Source:HGNC Symbol;Acc:HGNC:10637] |
| ENSG00000169248 | CXCL11 | C-X-C motif chemokine ligand 11 [Source:HGNC Symbol;Acc:HGNC:10638] |
| ENSG00000171016 | PYGO1 | pygopus family PHD finger 1 [Source:HGNC Symbol;Acc:HGNC:30256] |
| ENSG00000174807 | CD248 | CD248 molecule [Source:HGNC Symbol;Acc:HGNC:18219] |

*Table S3 continued from previous page*
| ensembl_gene_id | hgnc_symbol | description |
| --- | --- | --- |
| ENSG00000175318 | GRAMD2A | GRAM domain containing 2A [Source:HGNC Symbol;Acc:HGNC:27287] |
| ENSG00000175745 | NR2F1 | nuclear receptor subfamily 2 group F member 1 [Source:HGNC Symbol;Acc:HGNC:7975] |
| ENSG00000176771 | NCKAP5 | NCK associated protein 5 [Source:HGNC Symbol;Acc:HGNC:29847] |
| ENSG00000176884 | GRIN1 | glutamate ionotropic receptor NMDA type subunit 1 [Source:HGNC Symbol;Acc:HGNC:4584] |
| ENSG00000177721 | ANXA2R | annexin A2 receptor [Source:HGNC Symbol;Acc:HGNC:33463] |
| ENSG00000178662 | CSRNP3 | cysteine and serine rich nuclear protein 3 [Source:HGNC Symbol;Acc:HGNC:30729] |
| ENSG00000182759 | MAFA | MAF bZIP transcription factor A [Source:HGNC Symbol;Acc:HGNC:23145] |
| ENSG00000183770 | FOXL2 | forkhead box L2 [Source:HGNC Symbol;Acc:HGNC:1092] |
| ENSG00000184937 | WT1 | WT1 transcription factor [Source:HGNC Symbol;Acc:HGNC:12796] |
| ENSG00000184979 | USP18 | ubiquitin specific peptidase 18 [Source:HGNC Symbol;Acc:HGNC:12616] |
| ENSG00000185745 | IFIT1 | interferon induced protein with tetratricopeptide repeats 1 [Source:HGNC Symbol;Acc:HGNC:5407] |
| ENSG00000187608 | ISG15 | ISG15 ubiquitin like modifier [Source:HGNC Symbol;Acc:HGNC:4053] |
| ENSG00000189056 | RELN | reelin [Source:HGNC Symbol;Acc:HGNC:9957] |
| ENSG00000196159 | FAT4 | FAT atypical cadherin 4 [Source:HGNC Symbol;Acc:HGNC:23109] |
| ENSG00000198673 | TAFA2 | TAFA chemokine like family member 2 [Source:HGNC Symbol;Acc:HGNC:21589] |
| ENSG00000198963 | RORB | RAR related orphan receptor B [Source:HGNC Symbol;Acc:HGNC:10259] |
| ENSG00000203930 | LINC00632 | long intergenic non-protein coding RNA 632 [Source:HGNC Symbol;Acc:HGNC:27865] |
| ENSG00000204262 | COL5A2 | collagen type V alpha 2 chain [Source:HGNC Symbol;Acc:HGNC:2210] |
| ENSG00000223387 | LINC02068 | long intergenic non-protein coding RNA 2068 [Source:HGNC Symbol;Acc:HGNC:52914] |
| ENSG00000225964 | NRIR | negative regulator of interferon response [Source:HGNC Symbol;Acc:HGNC:51269] |

*Table S3 continued from previous page*
| ensembl_gene_id | hgnc_symbol | description |
| --- | --- | --- |
| ENSG00000226004 | SIMALR | suppressor of inflammatory macrophage apoptosis lncRNA [Source:HGNC Symbol;Acc:HGNC:53554] |
| ENSG00000231528 | FAM225A | family with sequence similarity 225 member A [Source:HGNC Symbol;Acc:HGNC:27855] |
| ENSG00000233608 | TWIST2 | twist family bHLH transcription factor 2 [Source:HGNC Symbol;Acc:HGNC:20670] |
| ENSG00000233975 | LINC02574 | long intergenic non-protein coding RNA 2574 [Source:HGNC Symbol;Acc:HGNC:53746] |
| ENSG00000239556 | UPK3BL3P | uroplakin 3B like 3, pseudogene [Source:HGNC Symbol;Acc:HGNC:55757] |
| ENSG00000259030 | FPGT-TNNI3K | FPGT-TNNI3K readthrough [Source:HGNC Symbol;Acc:HGNC:42952] |
| ENSG00000259275 |  | novel transcript |
| ENSG00000278599 | TBC1D3E | TBC1 domain family member 3E [Source:HGNC Symbol;Acc:HGNC:27071] |
| ENSG00000289445 |  | novel transcript |
| ENSG00000290525 | GBP1P1 | guanylate binding protein 1 pseudogene 1 [Source:HGNC Symbol;Acc:HGNC:39561] |
| ENSG00000299483 |  | novel transcript, antisense to LIPA |
| ENSG00000302633 |  | novel transcript |

**Table S4:** Annotation of the 9 input Ensembl genes across HPV16 vs non-HPV16 infections.

| ensembl_gene_id | hgnc_symbol | description |
| --- | --- | --- |
| ENSG00000145934 | TENM2 | teneurin transmembrane protein 2<br>[Source:HGNC Symbol;Acc:HGNC:29943] |
| ENSG00000178662 | CSRNP3 | cysteine and serine rich nuclear protein 3<br>[Source:HGNC Symbol;Acc:HGNC:30729] |
| ENSG00000186844 | LCE1A | late cornified envelope 1A [Source:HGNC Symbol;Acc:HGNC:29459] |
| ENSG00000210082 | MT-RNR2 | mitochondrially encoded 16S rRNA<br>[Source:HGNC Symbol;Acc:HGNC:7471] |
| ENSG00000211459 | MT-RNR1 | mitochondrially encoded 12S rRNA<br>[Source:HGNC Symbol;Acc:HGNC:7470] |
| ENSG00000251634 | NAIPP4 | NAIP pseudogene 4 [Source:HGNC Symbol;Acc:HGNC:54494] |
| ENSG00000274917 | RNA5-8SN5 | RNA, 5.8S ribosomal N5 [Source:HGNC Symbol;Acc:HGNC:53533] |
| ENSG00000278189 | RNA5-8SN1 | RNA, 5.8S ribosomal N1 [Source:HGNC Symbol;Acc:HGNC:53517] |
| ENSG00000285517 | LINC00941 | long intergenic non-protein coding RNA 941<br>[Source:HGNC Symbol;Acc:HGNC:48635] |

**Table S5:** Annotation of the 17 input Ensembl genes across cleared vs non-cleared infections.

| ensembl_gene_id | hgnc_symbol | description |
| --- | --- | --- |
| ENSG00000180658 | OR2A4 | olfactory receptor family 2 subfamily A member 4 [Source:HGNC Symbol;Acc:HGNC:14729] |
| ENSG00000186844 | LCE1A | late cornified envelope 1A [Source:HGNC Symbol;Acc:HGNC:29459] |
| ENSG00000187180 | LCE2C | late cornified envelope 2C [Source:HGNC Symbol;Acc:HGNC:29460] |
| ENSG00000198886 | MT-ND4 | mitochondrially encoded NADH:ubiquinone oxidoreductase core subunit 4 [Source:HGNC Symbol;Acc:HGNC:7459] |
| ENSG00000198888 | MT-ND1 | mitochondrially encoded NADH:ubiquinone oxidoreductase core subunit 1 [Source:HGNC Symbol;Acc:HGNC:7455] |
| ENSG00000198899 | MT-ATP6 | mitochondrially encoded ATP synthase membrane subunit 6 [Source:HGNC Symbol;Acc:HGNC:7414] |
| ENSG00000210082 | MT-RNR2 | mitochondrially encoded 16S rRNA [Source:HGNC Symbol;Acc:HGNC:7471] |
| ENSG00000211459 | MT-RNR1 | mitochondrially encoded 12S rRNA [Source:HGNC Symbol;Acc:HGNC:7470] |
| ENSG00000211659 | IGLV3-25 | immunoglobulin lambda variable 3-25 [Source:HGNC Symbol;Acc:HGNC:5908] |
| ENSG00000224689 | ZNF812P | zinc finger protein 812, pseudogene [Source:HGNC Symbol;Acc:HGNC:33242] |
| ENSG00000228253 | MT-ATP8 | mitochondrially encoded ATP synthase membrane subunit 8 [Source:HGNC Symbol;Acc:HGNC:7415] |
| ENSG00000233954 | UQCRHL | ubiquinol-cytochrome c reductase hinge protein like [Source:HGNC Symbol;Acc:HGNC:51714] |
| ENSG00000237973 | MTCO1P12 | MT-CO1 pseudogene 12 [Source:HGNC Symbol;Acc:HGNC:52014] |
| ENSG00000274286 | ADRA2B | adrenoceptor alpha 2B [Source:HGNC Symbol;Acc:HGNC:282] |
| ENSG00000281453 | TGFB2-OT1 | TGFB2 overlapping transcript 1 [Source:HGNC Symbol;Acc:HGNC:50629] |
| ENSG00000295133 |  | novel transcript |
| ENSG00000301729 |  | novel transcript, antisense to GLRA3 |

**Table S6:**
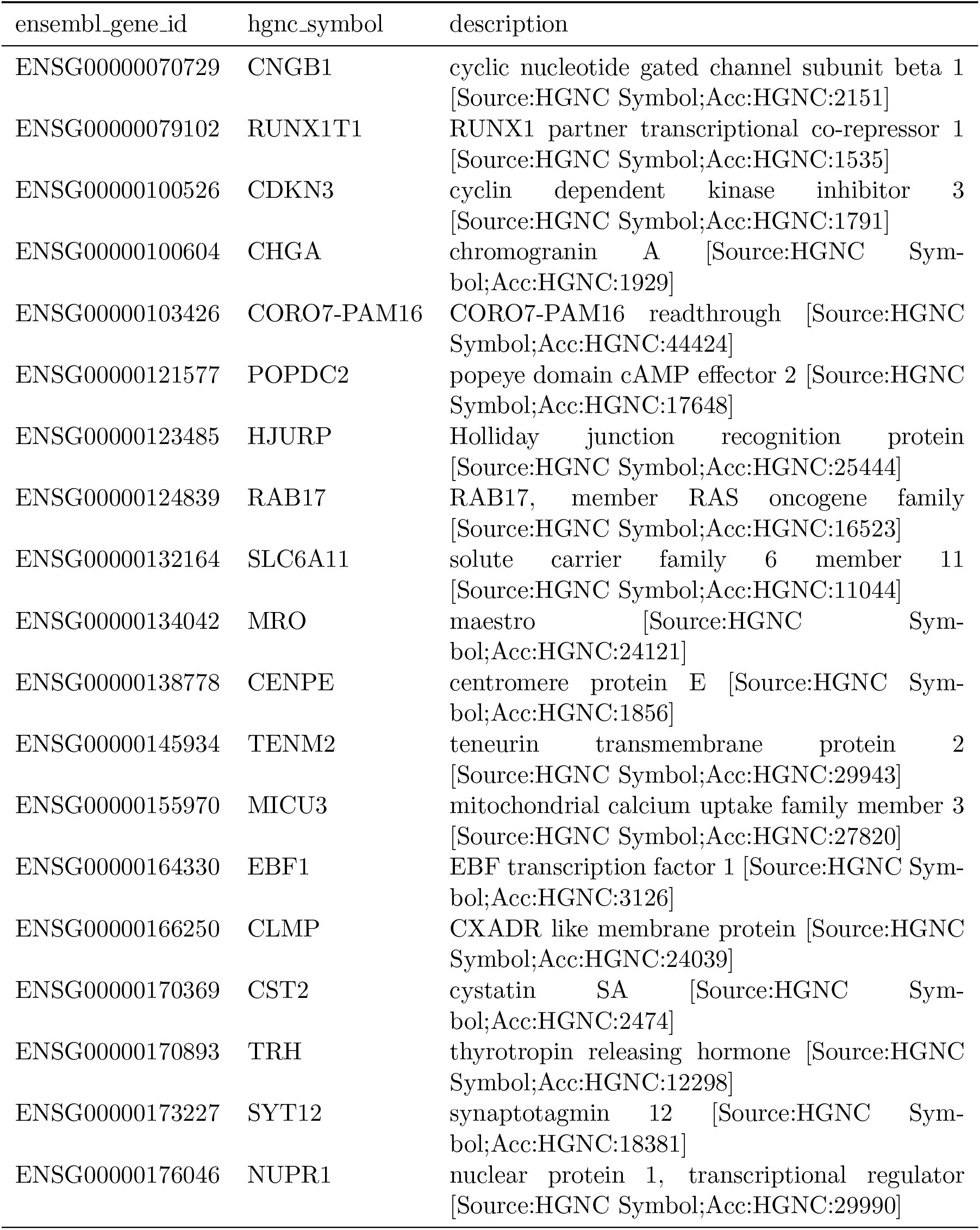
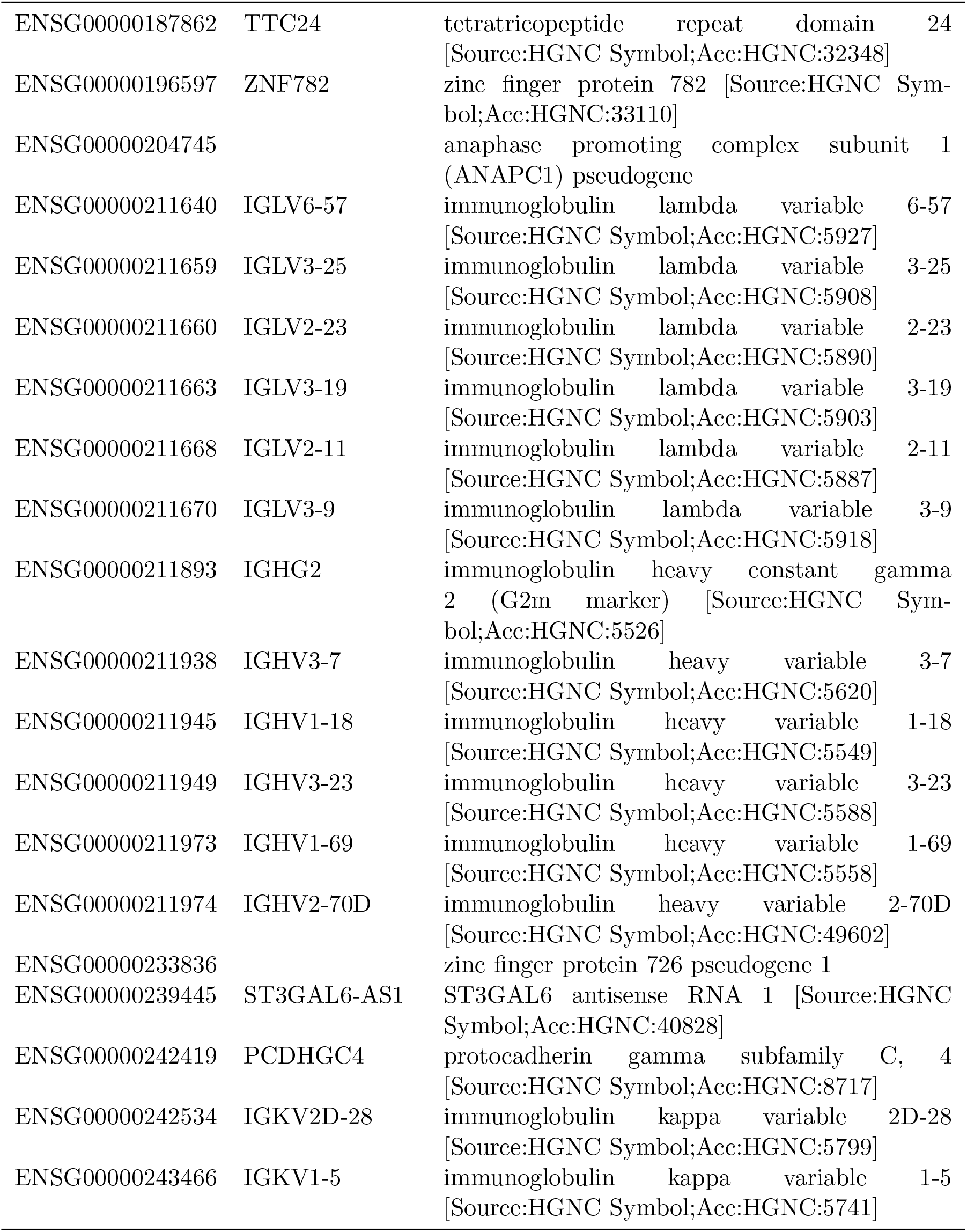
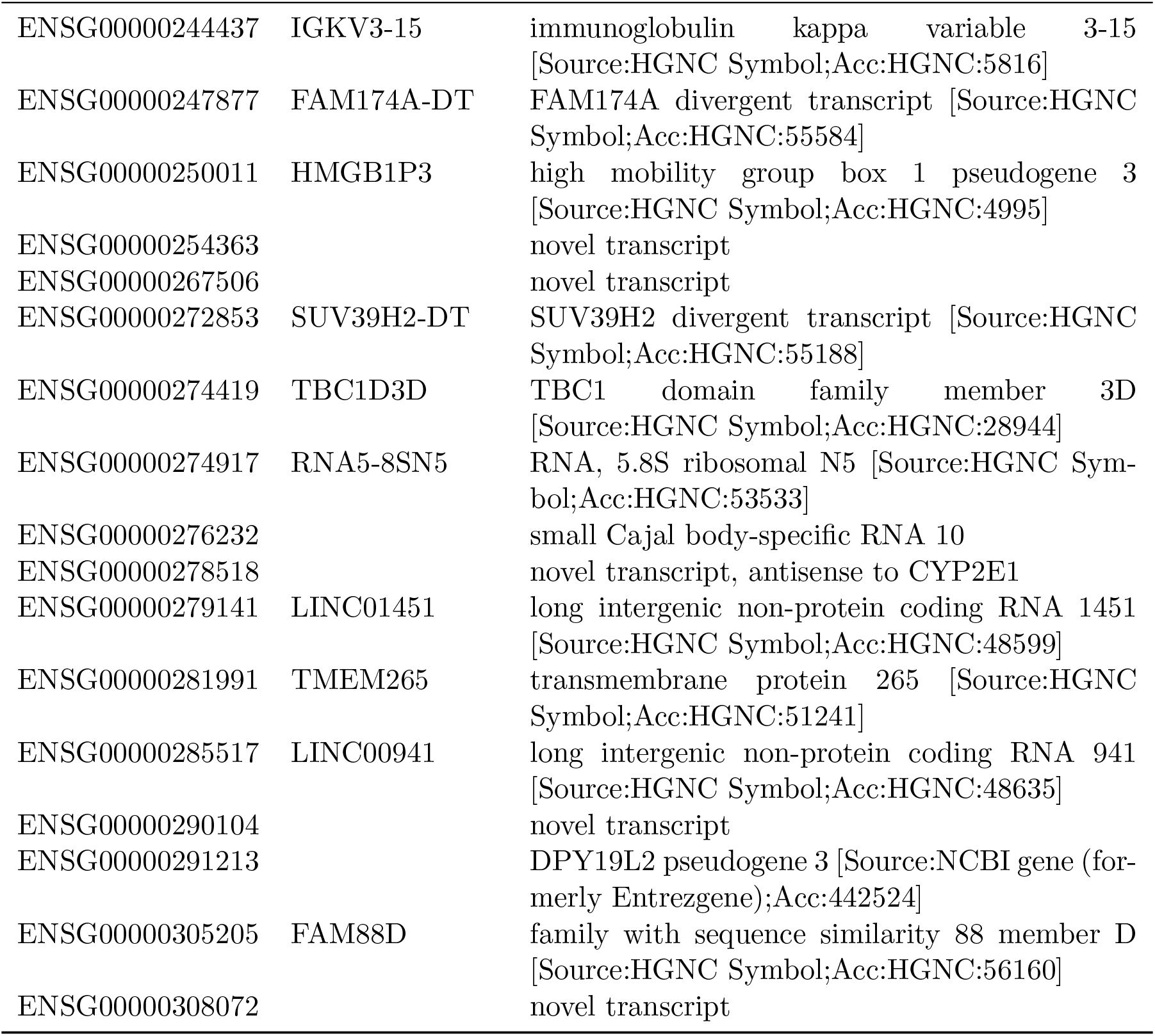
Annotation of the 56 input Ensembl genes across long, non-cleared vs the short, non-cleared infections.

**Figure S1:**
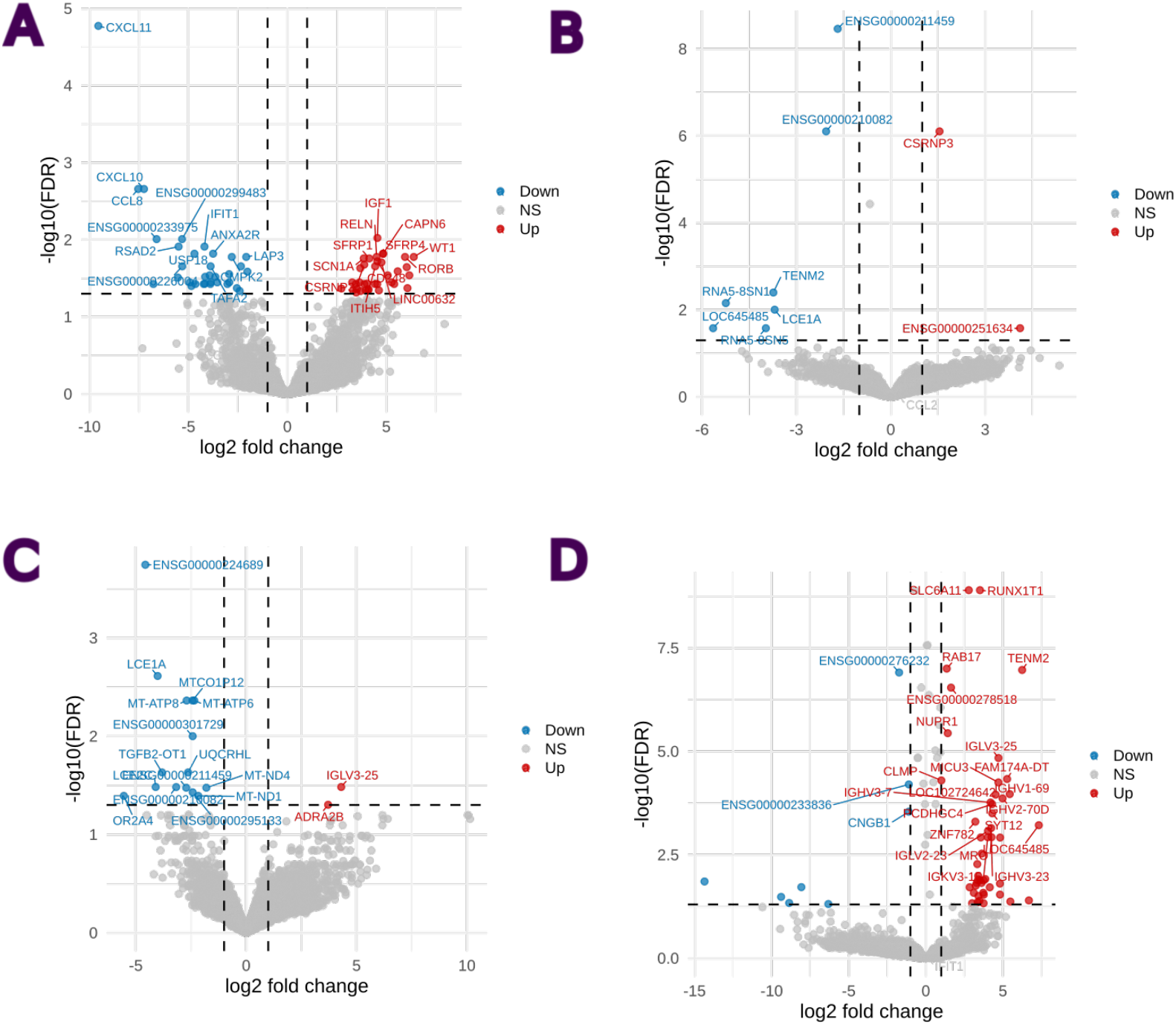
Differential gene expression across four comparisons: (A) *n* = 57 HPV-positive samples *vs*. *n* = 14 HPV-negative samples; (B) *n* = 26 HPV16 infections *vs*. *n* = 31 non-HPV16 infections; (C) *n* = 30 samples from non-clearing infections *vs*. *n* = 27 samples from clearing infections; and (D) *n* = 7 samples from long non-clearing infections *vs*. *n* = 23 samples from short non-clearing infections. Positive fold changes indicate higher expression and negative fold changes indicate lower expression. See Fig. 1 in the main text for details.

**Figure S2:**
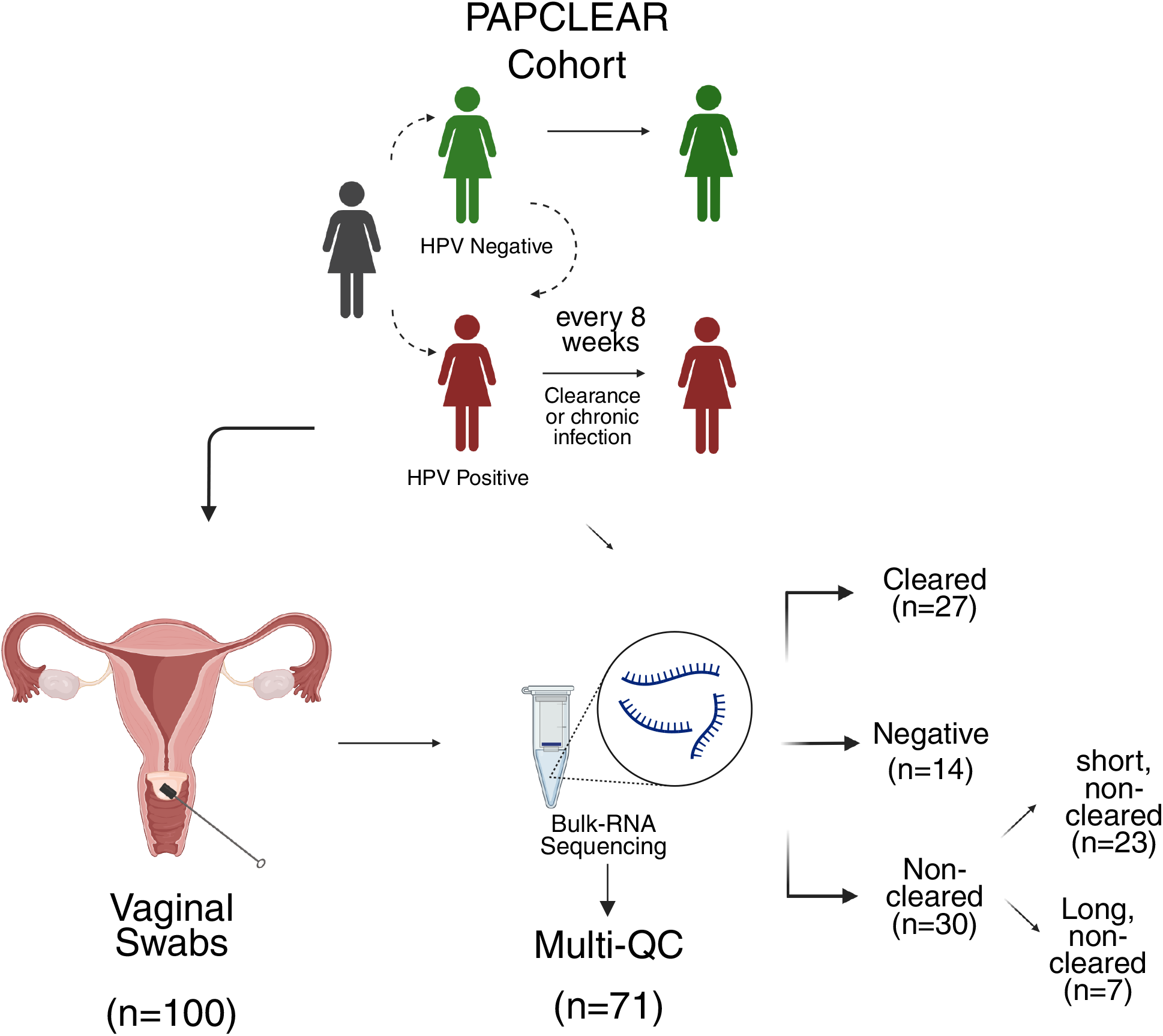
PAPCLEAR samples selection for bulk RNA sequencing. Infection clearance was defined by the absence of detection of the focal genotype at two consecutive visits. Participants who always tested negative or had only a transient HPV-positive sample were classified as ‘uninfected’. The infection outcome was further refined by distinguishing between long non-clearing *vs*. short non-clearing.

**Figure S3:**
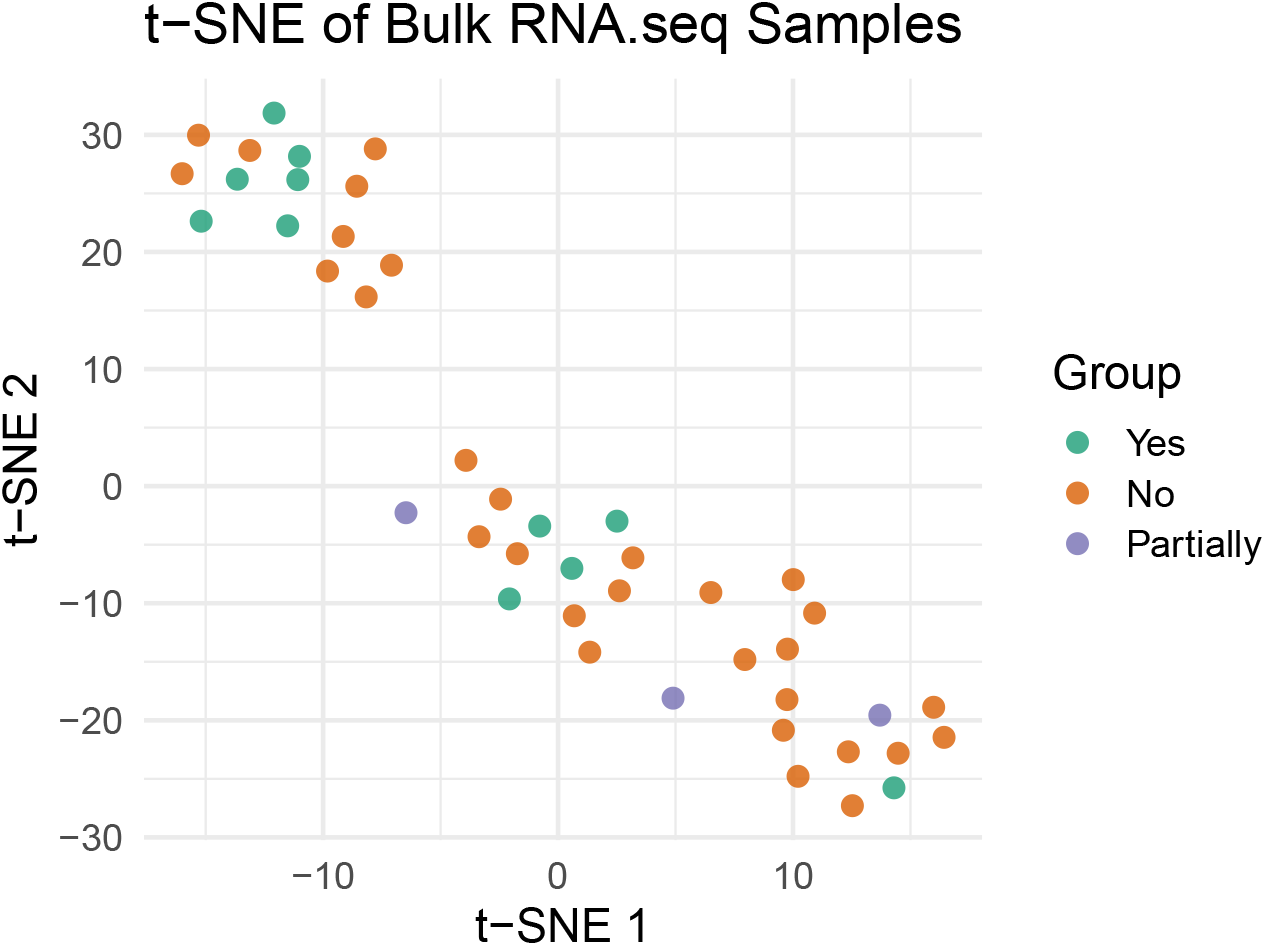
t-SNE clustering on the RNAseq data. Colours indicate the group to which the samples belong with regards to infection clearance.

**Figure S4:**
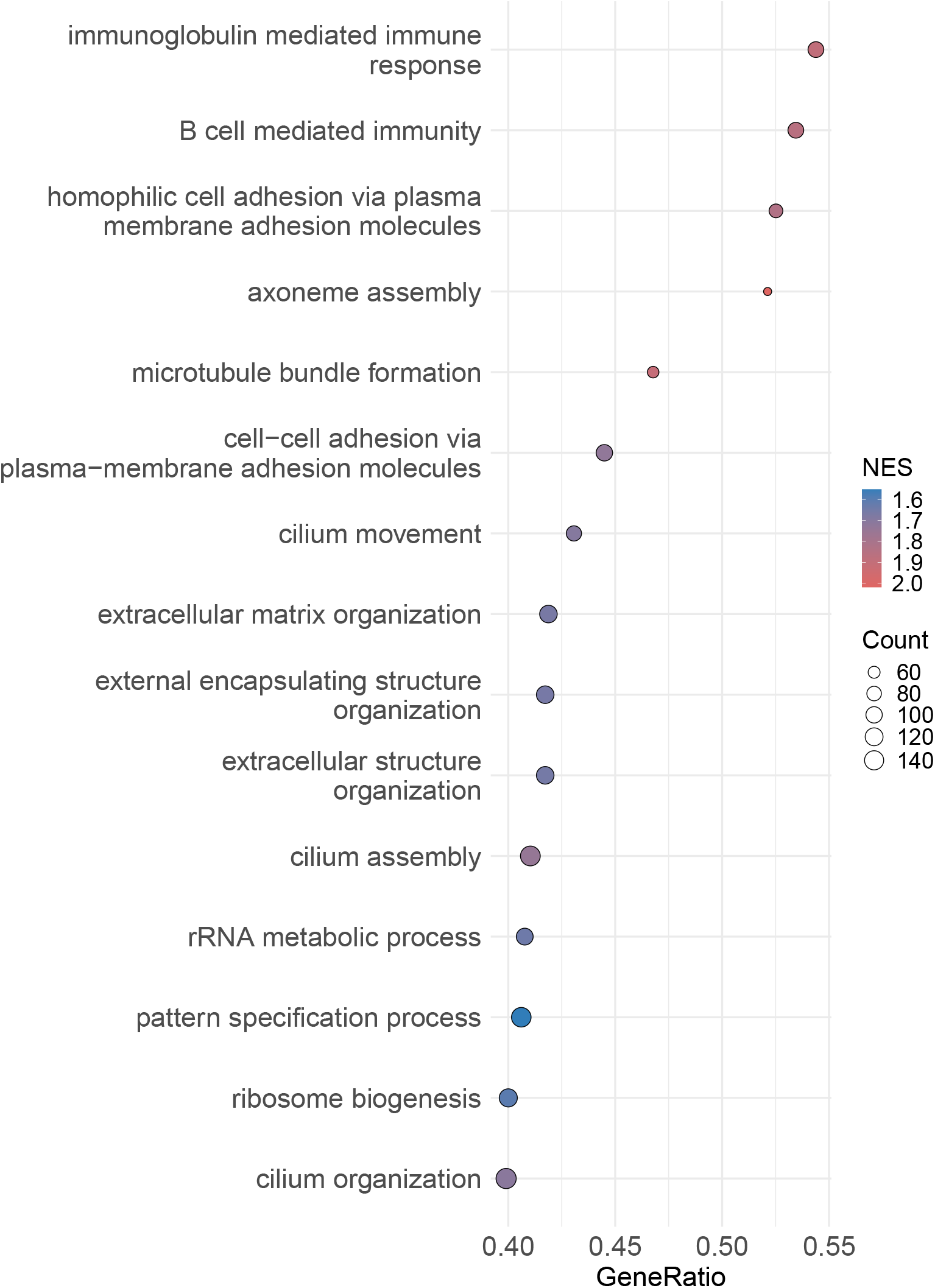
Dotplot of enriched pathways in non-clearing *vs*. clearing infections when only selecting a single sample per participant.

**Figure S5:**
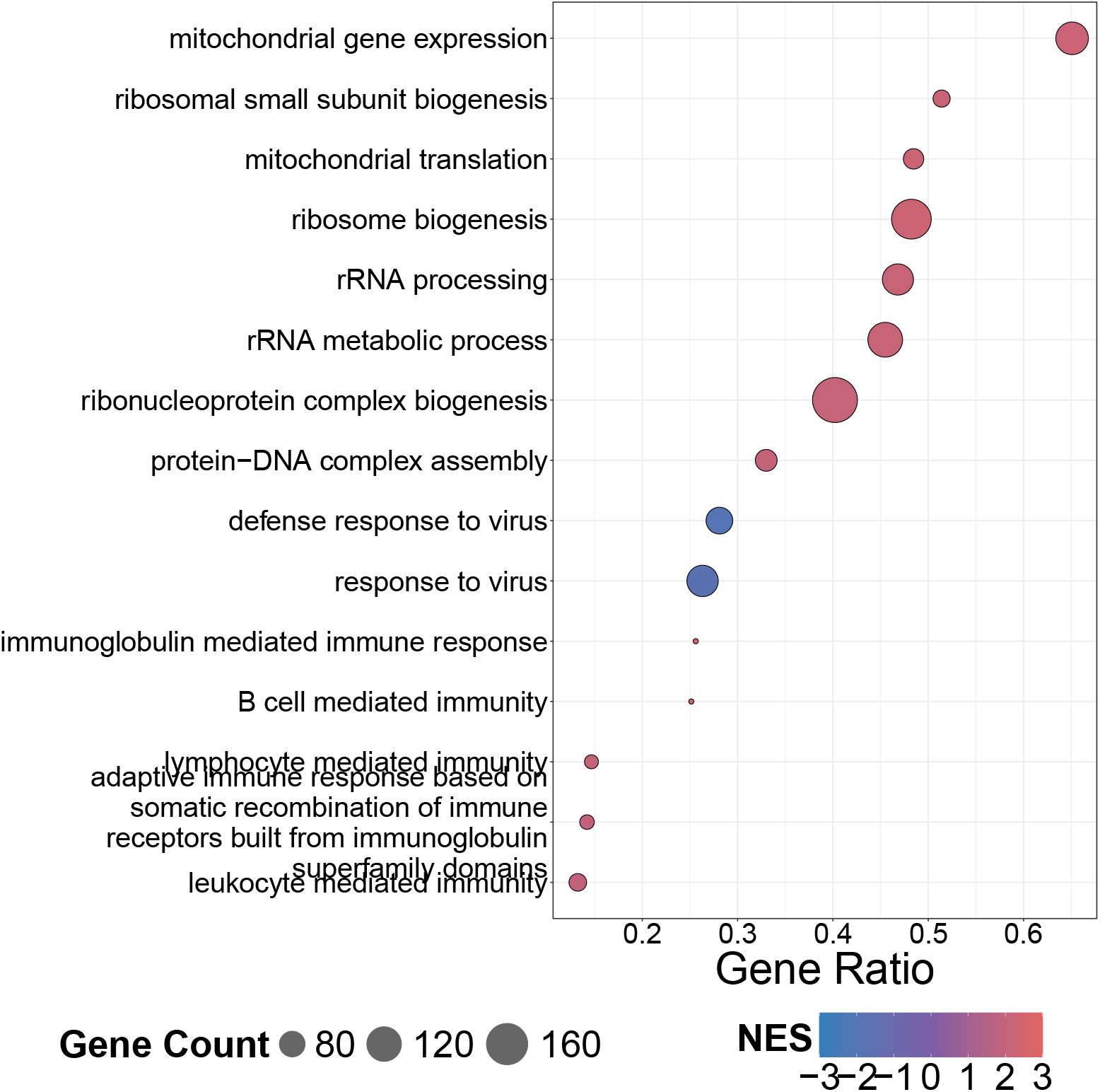
Dotplot of significantly expressed pathways in non-clearing infections compared to uninfected participants, ordered by gene-ratio.

**Figure S6:**
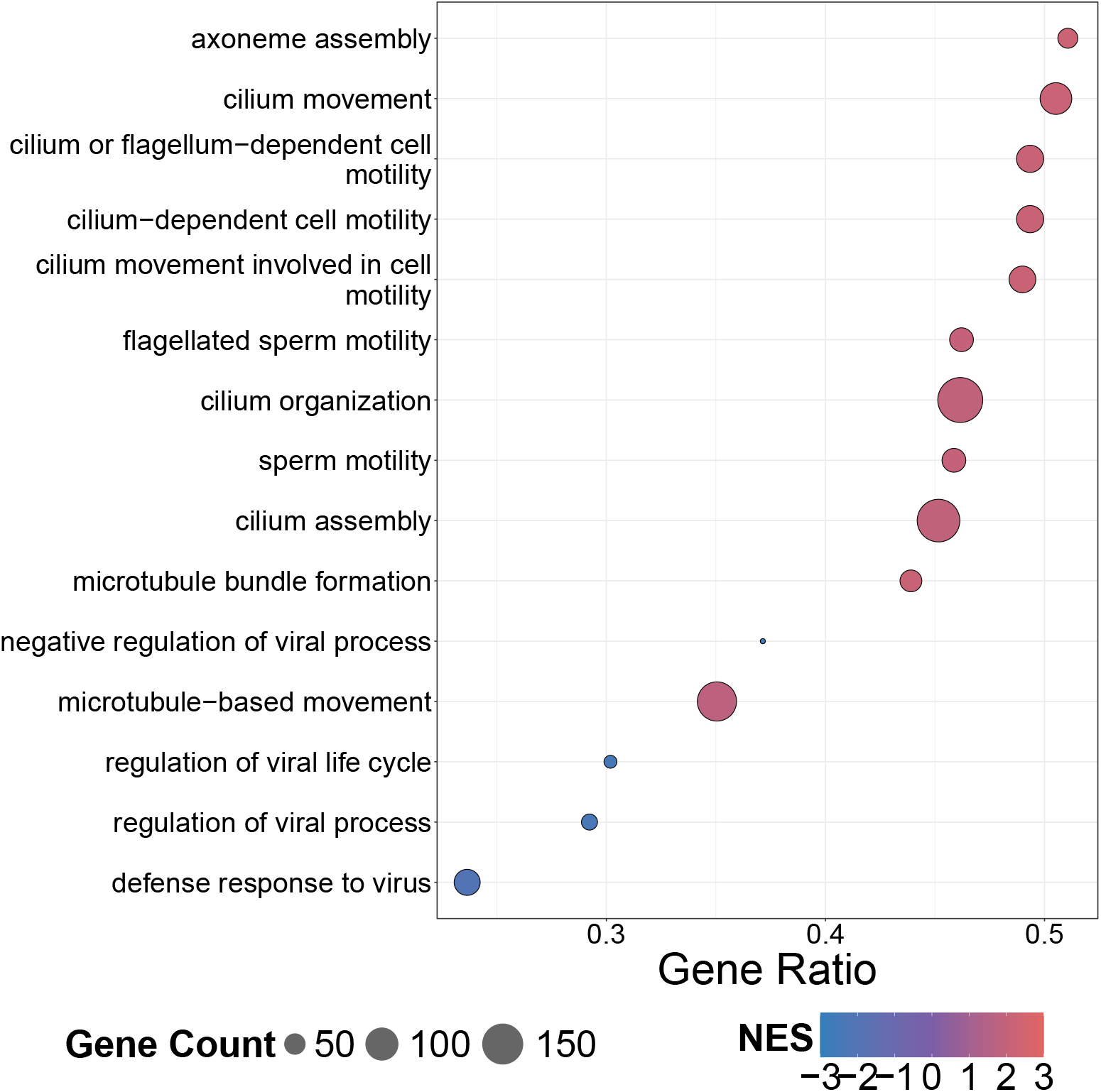
Dotplot of significantly expressed pathways in HPV16 infections compared to uninfected hosts.

